# Clonal Hematopoiesis and the Development and Progression of Metabolic Dysfunction-Associated Steatotic Liver Disease

**DOI:** 10.64898/2026.04.13.26350754

**Authors:** Ruijie Xie, Ben Schöttker

**Author notes:** **Corresponding author:** Prof. Dr. Ben Schöttker, PhD, Division of Clinical Epidemiology of Early Cancer Detection, German Cancer Research Center (DKFZ), Im Neuenheimer Feld 581, 69120 Heidelberg, Germany.

## Abstract

**Background & Aims:** Clonal hematopoiesis of indeterminate potential (CHIP) has been linked to chronic liver disease progression, yet its role across the full spectrum of metabolic dysfunction-associated steatotic liver disease (MASLD), from its initial development to end-stage complications, remains unclear. We aimed to comprehensively investigate the association of CHIP and its major subtypes with both the incidence and progression of MASLD.

**Methods:** We conducted a prospective cohort study of 353,218 UK Biobank participants, stratified into a healthy cohort free of MASLD at baseline (Cohort 1; n=230,270) and a prevalent MASLD cohort (Cohort 2; n=122,948). CHIP was ascertained from whole-exome sequencing data. We used multivariable Cox regression, competing risk models, and mediation analyses to assess the associations of CHIP (overall, by driver gene, and by clone size) with incident MASLD, cirrhosis, hepatocellular carcinoma (HCC), and liver-related death.

**Results:** In Cohort 1, CHIP was associated with an increased risk of incident MASLD (HR 1.25, 95% CI 1.08-1.44) and cirrhosis (HR 1.57, 95% CI 1.10-2.25). These associations were driven by non-DNMT3A mutations, particularly TET2, and showed a linear dose-response relationship with clone size. In Cohort 2, non-DNMT3A CHIP was associated with progression to cirrhosis (HR 1.82, 95% CI 1.28-2.58). The associations were more pronounced in males and in individuals without obesity or diabetes. C-reactive protein partially mediated the CHIP-MASLD association.

**Conclusion:** CHIP, driven predominantly by non-DNMT3A mutations (particularly TET2) is an independent risk factor for both the development and progression of MASLD. These findings position CHIP as a novel player in the pathophysiology of MASLD and suggest potential avenues for risk stratification and targeted anti-inflammatory intervention.

**Impact and Implications:** This large-scale, prospective study establishes clonal hematopoiesis of indeterminate potential (CHIP) as a novel and independent risk factor for the entire spectrum of metabolic dysfunction-associated steatotic liver disease (MASLD), from its initial development to its progression to cirrhosis and liver-related death. For hepatologists and hematologists, these findings identify a genetically defined, high-risk subpopulation, particularly individuals with non-DNMT3A mutations, who may benefit from enhanced liver surveillance. The identification of systemic inflammation as a partial mediator of the CHIP-MASLD association suggests that anti-inflammatory therapies currently under development for liver disease could represent a targeted treatment strategy for this growing patient population.

## Introduction

Metabolic dysfunction-associated steatotic liver disease (MASLD), formerly non-alcoholic fatty liver disease (NAFLD), is the most common chronic liver disease worldwide, affecting approximately one-third of the adult population.^[1, 2]^ MASLD encompasses a spectrum of conditions ranging from simple hepatic steatosis to non-alcoholic steatohepatitis (NASH), which is characterized by hepatocellular injury and necroinflammation and can progress to fibrosis, cirrhosis, and hepatocellular carcinoma (HCC).^[2, 3]^ With the rising global prevalence of obesity and type 2 diabetes, MASLD has become the fastest-growing etiology for HCC and liver transplantation.^[2, 4]^ Although advanced fibrosis is the strongest predictor of liver-related morbidity and mortality, the molecular and cellular drivers that determine why only a subset of individuals progress to severe disease remain incompletely understood.^[3, 5]^

Clonal hematopoiesis of indeterminate potential (CHIP) is an age-related condition defined by the clonal expansion of hematopoietic stem cells harboring somatic mutations in leukemia-associated driver genes, in the absence of overt hematologic malignancy.^[6, 7]^ Its prevalence increases sharply with age, from less than 1% in individuals younger than 50 years to over 10% in those older than 70 years.^[8, 9]^ Beyond a modestly elevated risk for hematologic malignancies, CHIP has emerged as a potent risk factor for a range of non-malignant inflammatory diseases, most notably conferring a two-fold increased risk of atherosclerotic cardiovascular disease and all-cause mortality.^[8, 10]^ The most commonly mutated genes in CHIP are the epigenetic regulators DNMT3A, TET2, and ASXL1, each of which may exert distinct biological effects.^[8, 9]^

The pathogenic effects of CHIP are thought to be mediated by a chronic, low-grade inflammatory state driven by mutant immune cells, particularly monocytes and macrophages.^[11]^ Experimental studies have demonstrated that CHIP-driver mutations, especially in TET2, augment the production of pro-inflammatory cytokines such as interleukin-1β via the NLRP3 inflammasome.^[10, 12]^ Given that macrophage-mediated inflammation is a central pathogenic mechanism in the progression of MASLD to NASH and fibrosis,^[13, 14]^ a potential role for CHIP in liver disease has been proposed.

Recent studies have provided the first evidence supporting this hypothesis, associating CHIP with an increased risk of chronic liver disease and HCC.^[15, 16]^ Despite these advances, whether CHIP contributes to the initial development of MASLD and how specific driver mutations and clone sizes differentially affect disease progression across the full clinical spectrum remain unclear.

Using data from the UK Biobank, we aimed to comprehensively investigate the association of CHIP and its major subtypes with both the incidence and progression of liver disease. We assessed the impact of CHIP on the risk of incident MASLD in a healthy population, as well as on the risk of progression to cirrhosis, HCC, and liver-related death, exploring the effects of specific driver genes and clone sizes across the disease spectrum.

## Materials and Methods

### Study population

This prospective cohort study was based on data from the UK Biobank, a population-based cohort that enrolled over 500,000 participants aged 40 to 69 years at 22 assessment centers across the United Kingdom between 2006 and 2010.^[17]^ At baseline, participants underwent a comprehensive assessment that included a self-administered touchscreen questionnaire, a verbal interview, physical measurements, and biological sample collection. The UK Biobank study was approved by the North West Multi-centre Research Ethics Committee (reference 11/NW/0382), and all participants provided written informed consent. The present analyses were conducted under UK Biobank application number 101633.

Of the 501,936 participants enrolled in the UK Biobank, we sequentially excluded participants based on criteria detailed in **Supplementary Table S1**, including heavy alcohol consumption (n = 106,551), prevalent viral hepatitis (n = 538), toxic liver disease (n = 52), and advanced liver disease at baseline (n = 974). After also excluding those with unavailable CHIP data (n = 40,603), a final analytical cohort of 353,218 participants remained (**Figure 1**).

**Figure 1.**
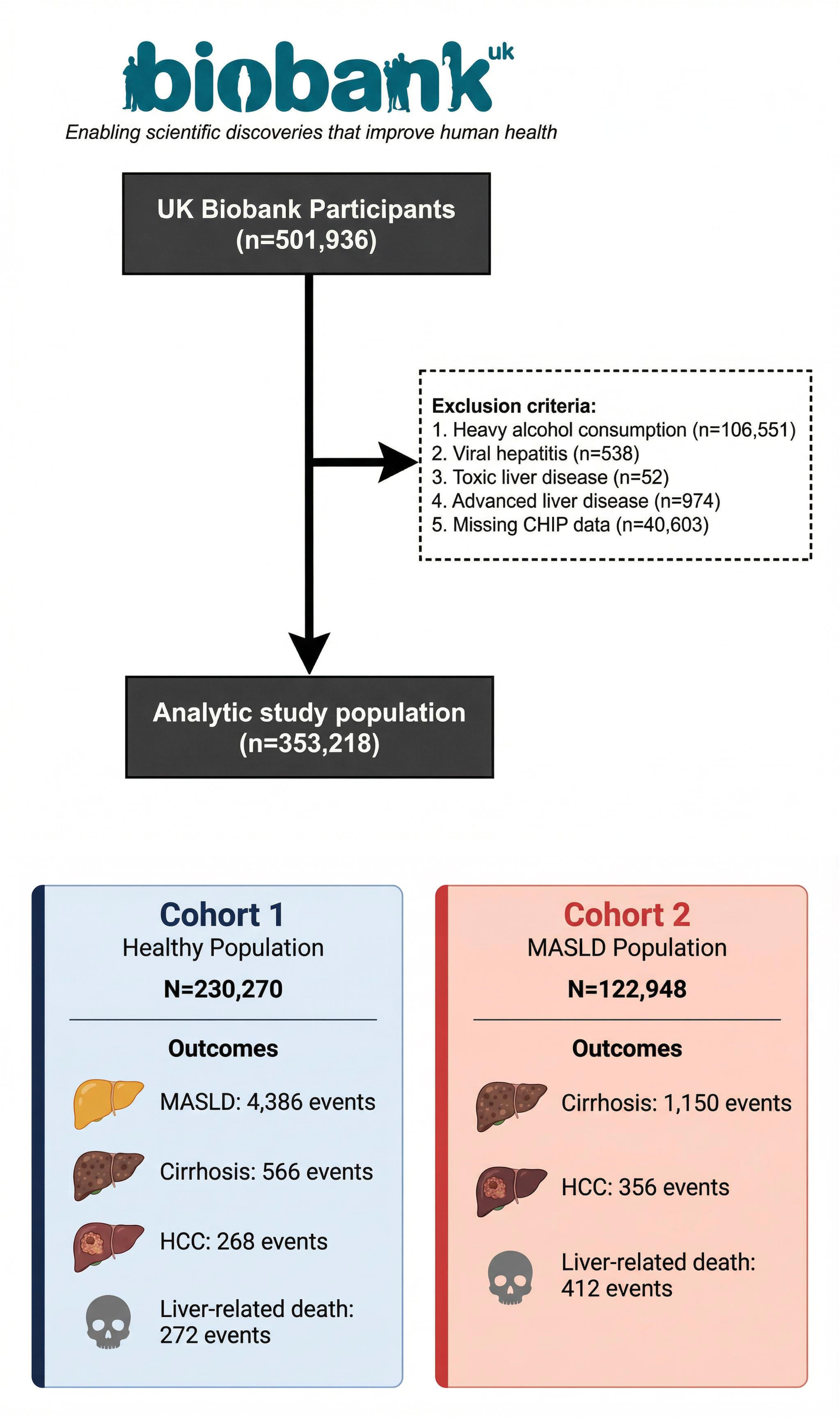
Study Design and Participant Flow **Abbreviations:** CHIP, clonal hematopoiesis of indeterminate potential; MASLD, metabolic dysfunction-associated steatotic liver disease.

The analytical cohort was then stratified into two groups based on baseline MASLD status. Cohort 1 (Healthy Population; N = 230,270) included participants free of MASLD at baseline and was used to investigate the association of CHIP with the risk of incident MASLD and subsequent liver disease progression. Cohort 2 (MASLD Population; N = 122,948) comprised participants with prevalent MASLD at baseline and was used to evaluate the association of CHIP with the risk of disease progression to cirrhosis, HCC, and liver-related death.

### Definition of MASLD and Outcomes

Baseline MASLD was ascertained using the Fatty Liver Index (FLI), a well-validated algorithm for the non-invasive prediction of hepatic steatosis, computed from body mass index (BMI), waist circumference, serum triglycerides, and gamma-glutamyl transferase (GGT).^[18]^ An FLI ≥60 was used to define the presence of hepatic steatosis, in accordance with prior UK Biobank studies and established validation data.^[19, 20]^ To fulfill the updated MASLD nomenclature criteria,^[1]^ participants with hepatic steatosis (FLI ≥60) were additionally required to have at least one cardiometabolic risk factor, as detailed in **Supplementary Table S2**.

Incident clinical outcomes were ascertained through algorithmic linkage to national electronic health records, including Hospital Episode Statistics (HES) for England, Scottish Morbidity Records (SMR), and the Patient Episode Database for Wales (PEDW), as well as national death and cancer registries. The follow-up period commenced at the date of baseline assessment and extended until the date of first event, death, loss to follow-up, or the administrative censoring date (March 31, 2022), whichever occurred first. Detailed definitions for all clinical outcomes, including the specific International Classification of Diseases, 10th Revision (ICD-10) codes used, are provided in **Supplementary Table S3**.

### Ascertainment of CHIP

CHIP status was obtained from the UK Biobank return dataset (Category 170; Data-Fields 30105-30107), which provides curated somatic mutation data for 450,679 participants. This dataset was derived from blood-derived whole-exome sequencing (WES) data processed by the UK Biobank consortium using a standardized and validated bioinformatics pipeline, as described by Vlasschaert et al.^[21]^ Briefly, somatic variant calling was performed using Mutect2 from the Genome Analysis Toolkit (GATK) in tumor-only mode, targeting a canonical list of 74 genes recurrently mutated in myeloid malignancies. Potential germline variants were flagged using population allele frequency data, and putative variants were filtered based on sequencing quality metrics and population-level artifact removal. CHIP was defined as the presence of at least one somatic mutation with a variant allele fraction (VAF) of ≥2%.

For our analyses, CHIP carriers were further classified by clone size into Small CHIP (2% ≤ VAF < 10%) and Large CHIP (VAF ≥10%), and by the mutated driver gene (*DNMT3A*, *TET2*, *ASXL1*). A composite “non-*DNMT3A* CHIP” category was defined to include all CHIP carriers with mutations in any gene other than *DNMT3A*, given the distinct biological and clinical implications of *DNMT3A* versus non-*DNMT3A* mutations.^[15, 16]^

### Assessment of Covariates

Baseline covariates were collected during the initial assessment visit. Sociodemographic factors included age, sex, and ethnicity. Lifestyle factors included education level, Townsend deprivation index, smoking status, physical activity, daily alcohol intake, a diet risk score, and body mass index (BMI). Clinical comorbidities included prevalent diabetes, hypertension, and statin use. Laboratory biomarkers included triglycerides, HDL cholesterol, C-reactive protein (CRP), alanine aminotransferase (ALT), GGT, and platelet count. The systemic immune-inflammation index (SII) was calculated as platelet count × neutrophil count / lymphocyte count.^[22]^ The Fibrosis-4 (FIB-4) index was calculated from age, aspartate aminotransferase (AST), ALT, and platelet count.^[23]^

Detailed definitions, coding, and measurement methods for all covariates are provided in **Supplementary Table S4.**

### Statistical Analysis

Baseline characteristics were summarized as means (standard deviation [SD]) or medians (interquartile range [IQR]) for continuous variables and as frequencies (percentages) for categorical variables. Between-group differences were assessed using independent-samples t-tests or Mann-Whitney U tests for continuous variables and chi-squared tests for categorical variables, as appropriate. The proportion of missing data across covariates ranged from 0% to 15.4% (glucose). Missing values were imputed using the missForest algorithm, a non-parametric iterative imputation method based on random forests.^[24]^

Multivariable Cox proportional hazards regression models were used to estimate hazard ratios (HRs) and 95% confidence intervals (CIs) for the associations of CHIP status and gene-specific subtypes with each liver outcome. We constructed three nested models with sequential covariate adjustment: **Model 1** adjusted for age, sex, and ethnicity; **Model 2** additionally adjusted for education level, Townsend deprivation index, smoking status, physical activity, daily alcohol intake, diet risk score, and BMI; **Model 3** (the fully adjusted model) further adjusted for prevalent diabetes, hypertension, statin use, and baseline levels of triglycerides, HDL cholesterol, CRP, ALT, GGT, and platelet count. To account for the competing risk of non-liver-related death, we additionally estimated subdistribution hazard ratios (SHRs) from the fully adjusted model using the Fine-Gray proportional subdistribution hazards model.^[25]^

To characterize the shape of the dose-response relationship between CHIP clone size and liver outcomes, we modeled VAF as a continuous variable using restricted cubic splines (RCS) with four knots. Non-linearity was assessed by comparing the model with the spline terms to a model with only a linear term using a likelihood ratio test. Pre-specified subgroup analyses were conducted to evaluate potential effect modification by age, sex, BMI, smoking status, prevalent diabetes, and ethnicity, with P-values for interaction derived from multiplicative interaction terms.

To explore potential biological mechanisms, we performed causal mediation analyses using the product-of-coefficients approach.^[26]^ The candidate mediators included inflammatory biomarkers (white blood cell count, neutrophil count, monocyte count, lymphocyte count, platelet count, CRP, SII), liver enzymes (ALT, AST, GGT), and a panel of 25 circulating cytokines. The proportion of the total effect mediated through each biomarker was estimated with 95% CIs obtained via bootstrapping (1,000 resamples).

Cross-sectional associations between CHIP and a panel of metabolic biomarkers at baseline were assessed in the total population using multivariable logistic regression, adjusted for the same set of covariates as Model 2. Detailed definitions for these biomarker cut-offs are provided in **Supplementary Table S5**.

All statistical analyses were performed using R software (version 4.2.0; R Foundation for Statistical Computing, Vienna, Austria). A two-sided P-value < 0.05 was considered statistically significant.

## Results

### Baseline characteristics of the study population

Of the 501,936 participants in the UK Biobank, our final analytical cohort comprised 353,218 individuals after applying exclusion criteria (**Figure 1**). This population was stratified into Cohort 1, including 230,270 participants free of MASLD at baseline (mean age 56.2 years, 66.4% female), and Cohort 2, including 122,948 participants with prevalent MASLD (mean age 57.4 years, 41.7% female). As detailed in **Table 1**, participants in Cohort 2 were, on average, older, more likely to be male, and had a significantly higher prevalence of cardiometabolic comorbidities, including diabetes, hypertension, and statin use, compared to those in Cohort 1. They also presented with a more adverse profile of liver enzymes, triglycerides, and inflammatory markers.

**Table 1.**
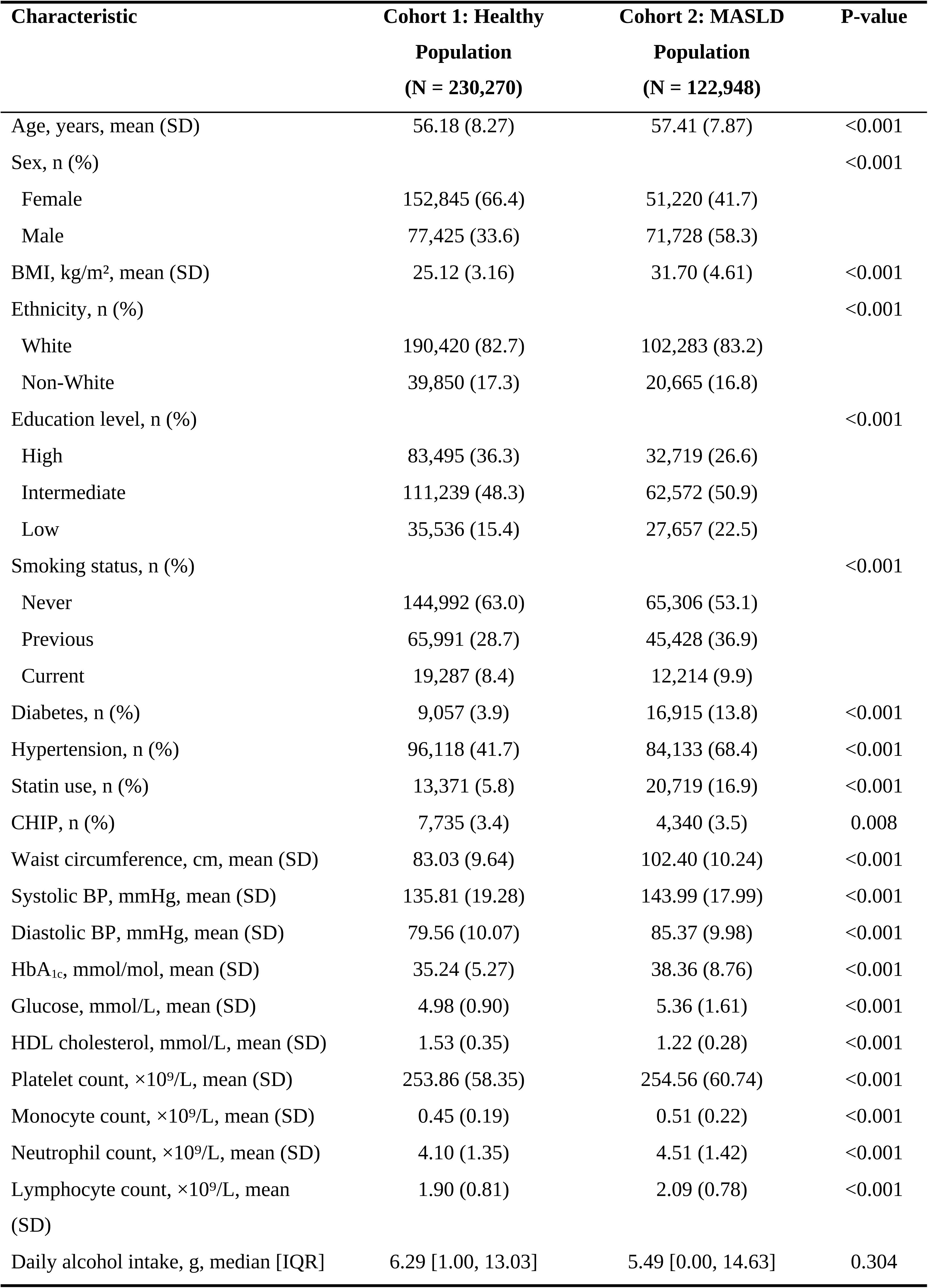
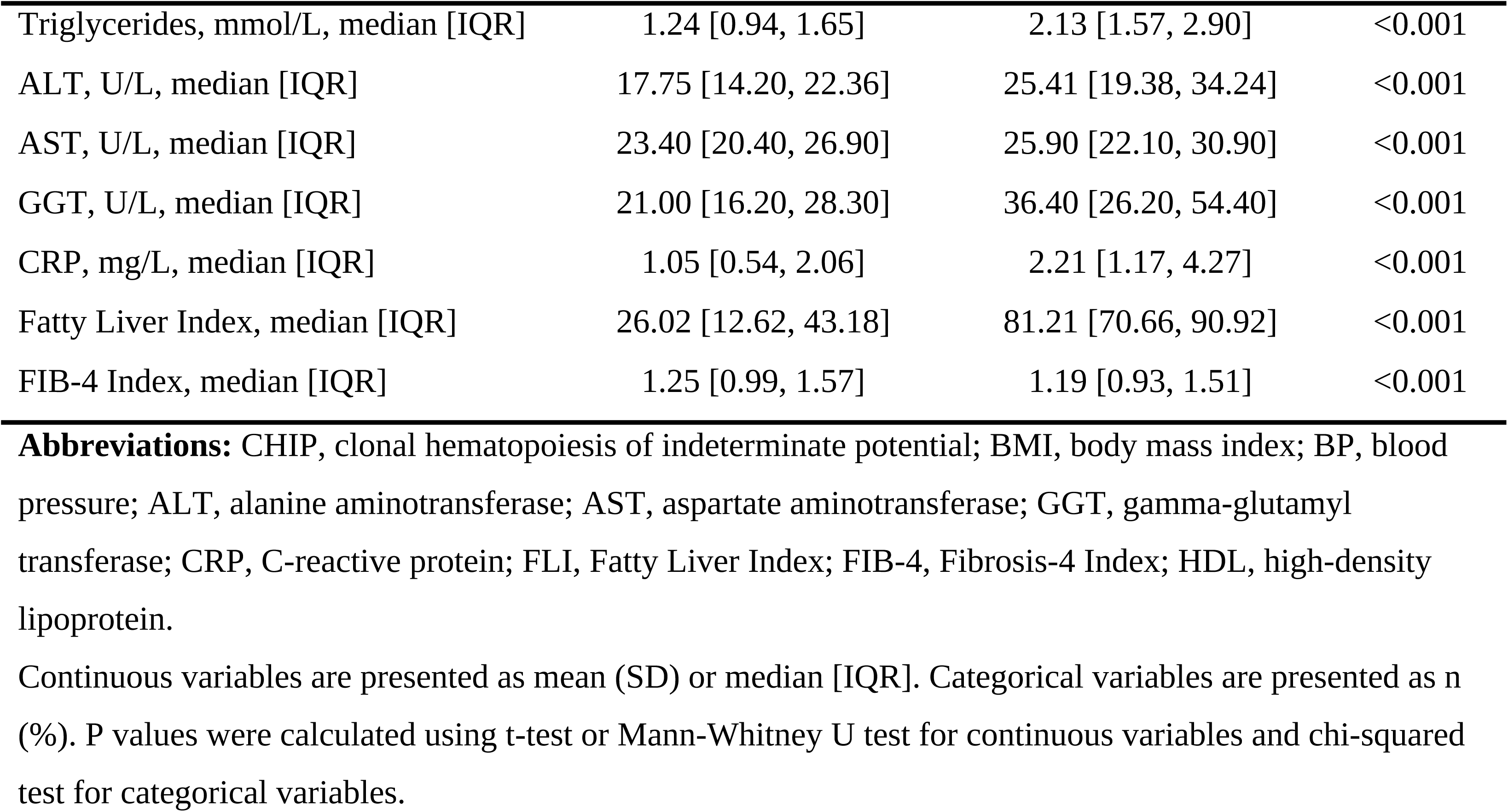
Baseline Characteristics of Study Participants.

| Characteristic | Cohort 1: Healthy<br>Population<br>(N = 230,270) | Cohort 2: MASLD<br>Population<br>(N = 122,948) | P-value |
| --- | --- | --- | --- |
| Age, years, mean (SD) | 56.18 (8.27) | 57.41 (7.87) | <0.001 |
| Sex, n (%) |  |  | <0.001 |
| Female | 152,845 (66.4) | 51,220 (41.7) |  |
| Male | 77,425 (33.6) | 71,728 (58.3) |  |
| BMI, kg/m<sup>2</sup>, mean (SD) | 25.12 (3.16) | 31.70 (4.61) | <0.001 |
| Ethnicity, n (%) |  |  | <0.001 |
| White | 190,420 (82.7) | 102,283 (83.2) |  |
| Non-White | 39,850 (17.3) | 20,665 (16.8) |  |
| Education level, n (%) |  |  | <0.001 |
| High | 83,495 (36.3) | 32,719 (26.6) |  |
| Intermediate | 111,239 (48.3) | 62,572 (50.9) |  |
| Low | 35,536 (15.4) | 27,657 (22.5) |  |
| Smoking status, n (%) |  |  | <0.001 |
| Never | 144,992 (63.0) | 65,306 (53.1) |  |
| Previous | 65,991 (28.7) | 45,428 (36.9) |  |
| Current | 19,287 (8.4) | 12,214 (9.9) |  |
| Diabetes, n (%) | 9,057 (3.9) | 16,915 (13.8) | <0.001 |
| Hypertension, n (%) | 96,118 (41.7) | 84,133 (68.4) | <0.001 |
| Statin use, n (%) | 13,371 (5.8) | 20,719 (16.9) | <0.001 |
| CHIP, n (%) | 7,735 (3.4) | 4,340 (3.5) | 0.008 |
| Waist circumference, cm, mean (SD) | 83.03 (9.64) | 102.40 (10.24) | <0.001 |
| Systolic BP, mmHg, mean (SD) | 135.81 (19.28) | 143.99 (17.99) | <0.001 |
| Diastolic BP, mmHg, mean (SD) | 79.56 (10.07) | 85.37 (9.98) | <0.001 |
| HbA<sub>1c</sub>, mmol/mol, mean (SD) | 35.24 (5.27) | 38.36 (8.76) | <0.001 |
| Glucose, mmol/L, mean (SD) | 4.98 (0.90) | 5.36 (1.61) | <0.001 |
| HDL cholesterol, mmol/L, mean (SD) | 1.53 (0.35) | 1.22 (0.28) | <0.001 |
| Platelet count, ×10<sup>9</sup>/L, mean (SD) | 253.86 (58.35) | 254.56 (60.74) | <0.001 |
| Monocyte count, ×10<sup>9</sup>/L, mean (SD) | 0.45 (0.19) | 0.51 (0.22) | <0.001 |
| Neutrophil count, ×10<sup>9</sup>/L, mean (SD) | 4.10 (1.35) | 4.51 (1.42) | <0.001 |
| Lymphocyte count, ×10<sup>9</sup>/L, mean<br>(SD) | 1.90 (0.81) | 2.09 (0.78) | <0.001 |
| Daily alcohol intake, g, median [IQR] | 6.29 [1.00, 13.03] | 5.49 [0.00, 14.63] | 0.304 |
| Triglycerides, mmol/L, median [IQR] | 1.24 [0.94, 1.65] | 2.13 [1.57, 2.90] | <0.001 |
| ALT, U/L, median [IQR] | 17.75 [14.20, 22.36] | 25.41 [19.38, 34.24] | <0.001 |
| AST, U/L, median [IQR] | 23.40 [20.40, 26.90] | 25.90 [22.10, 30.90] | <0.001 |
| GGT, U/L, median [IQR] | 21.00 [16.20, 28.30] | 36.40 [26.20, 54.40] | <0.001 |
| CRP, mg/L, median [IQR] | 1.05 [0.54, 2.06] | 2.21 [1.17, 4.27] | <0.001 |
| Fatty Liver Index, median [IQR] | 26.02 [12.62, 43.18] | 81.21 [70.66, 90.92] | <0.001 |
| FIB-4 Index, median [IQR] | 1.25 [0.99, 1.57] | 1.19 [0.93, 1.51] | <0.001 |

**Abbreviations:** CHIP, clonal hematopoiesis of indeterminate potential; BMI, body mass index; BP, blood pressure; ALT, alanine aminotransferase; AST, aspartate aminotransferase; GGT, gamma-glutamyl transferase; CRP, C-reactive protein; FLI, Fatty Liver Index; FIB-4, Fibrosis-4 Index; HDL, high-density lipoprotein.
Continuous variables are presented as mean (SD) or median [IQR]. Categorical variables are presented as n (%). P values were calculated using t-test or Mann-Whitney U test for continuous variables and chi-squared test for categorical variables.

CHIP was detected in 12,075 participants (3.4%) of the total study population, with a similar prevalence in Cohort 1 (7,735; 3.4%) and Cohort 2 (4,340; 3.5%; P=0.008). The prevalence of CHIP increased markedly with age, from <1% in participants younger than 50 years to >7% in those older than 70 years (**Supplementary Figure S1**). The most commonly mutated gene was DNMT3A (n=7,582; 62.8%), followed by TET2 (n=1,772; 14.7%) and ASXL1 (n=1,304; 10.8%).

### CHIP and risk of incident liver disease in the healthy population

During a median follow-up of 13.1 years, we documented 4,386 incident MASLD cases, 566 incident cirrhosis cases, 268 incident HCC cases, and 272 liver-related deaths in Cohort 1 (**Figure 2**). In the fully adjusted model, the presence of CHIP was associated with a 25% increased risk of incident MASLD (HR 1.25, 95% CI 1.08-1.44) and a 57% increased risk of incident cirrhosis (HR 1.57, 95% CI 1.10-2.25). Similar results were observed in Model 1 and Model 2 (**Supplementary Table S6**). This association was primarily driven by non-DNMT3A mutations, which conferred a 63% increased risk of incident MASLD (HR 1.63, 95% CI 1.35-1.98) and more than a two-fold increased risk of incident cirrhosis (HR 2.12, 95% CI 1.32-3.40) (**Figure 3**). Specifically, TET2-mutant CHIP was associated with a 72% higher risk of incident MASLD (HR 1.72, 95% CI 1.27-2.34) and a nearly three-fold increased risk of liver-related death (HR 2.78, 95% CI 1.14-6.75). A strong but non-significant trend was also observed for TET2-mutant CHIP and incident HCC (HR 2.18, 95% CI 0.81-5.86). By contrast, DNMT3A mutations alone were not significantly associated with any liver outcome in Cohort 1. When stratified by clone size, large CHIP (VAF ≥10%) was associated with a 38% higher risk of MASLD (HR 1.38, 95% CI 1.17-1.62) and a 66% higher risk of cirrhosis (HR 1.66, 95% CI 1.09-2.52). Restricted cubic spline analyses confirmed a positive, linear dose-response relationship between CHIP VAF and the risk of incident MASLD (P for non-linearity = 0.880) and cirrhosis (P for non-linearity = 0.978) (**Figure 4**). These findings remained consistent in competing risk analyses (**Supplementary Figure S2 and S3**).

**Figure 2.**
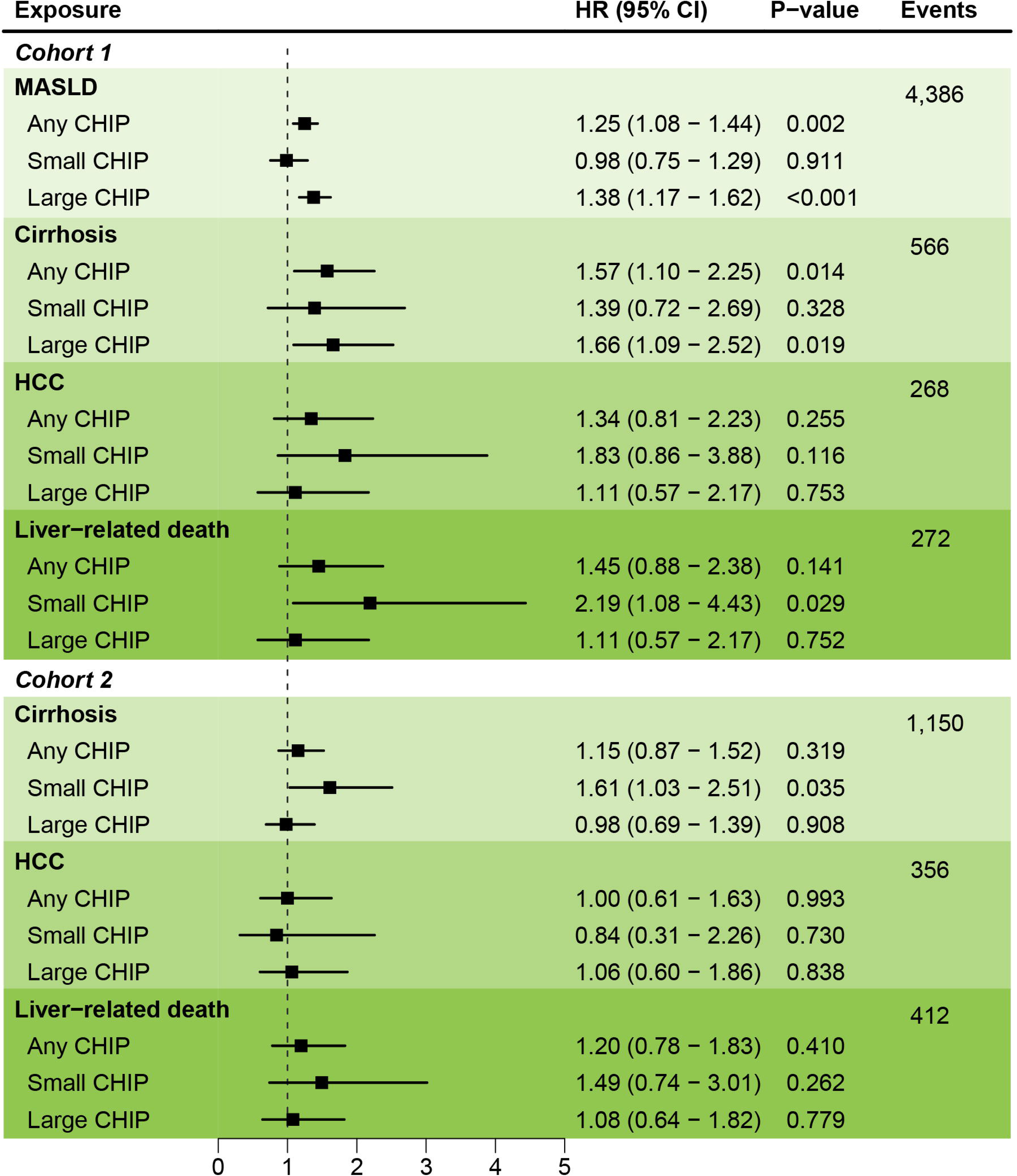
Association of CHIP (Overall and by Clone Size) with Liver Outcomes Forest plot showing HRs and 95% CIs for the associations of CHIP, stratified by presence (Any CHIP) and clone size (Small CHIP: 2% ≤ VAF < 10%; Large CHIP: VAF ≥10%), with liver-related outcomes in Cohort 1 (incident MASLD, cirrhosis, HCC, and liver-related death) and Cohort 2 (cirrhosis, HCC, and liver-related death). Results are from the fully adjusted Cox proportional hazards model (Model 3). **Abbreviations:** CHIP, clonal hematopoiesis of indeterminate potential; CI, confidence interval; HCC, hepatocellular carcinoma; HR, hazard ratio; MASLD, metabolic dysfunction-associated steatotic liver disease; VAF, variant allele fraction.

**Figure 3.**
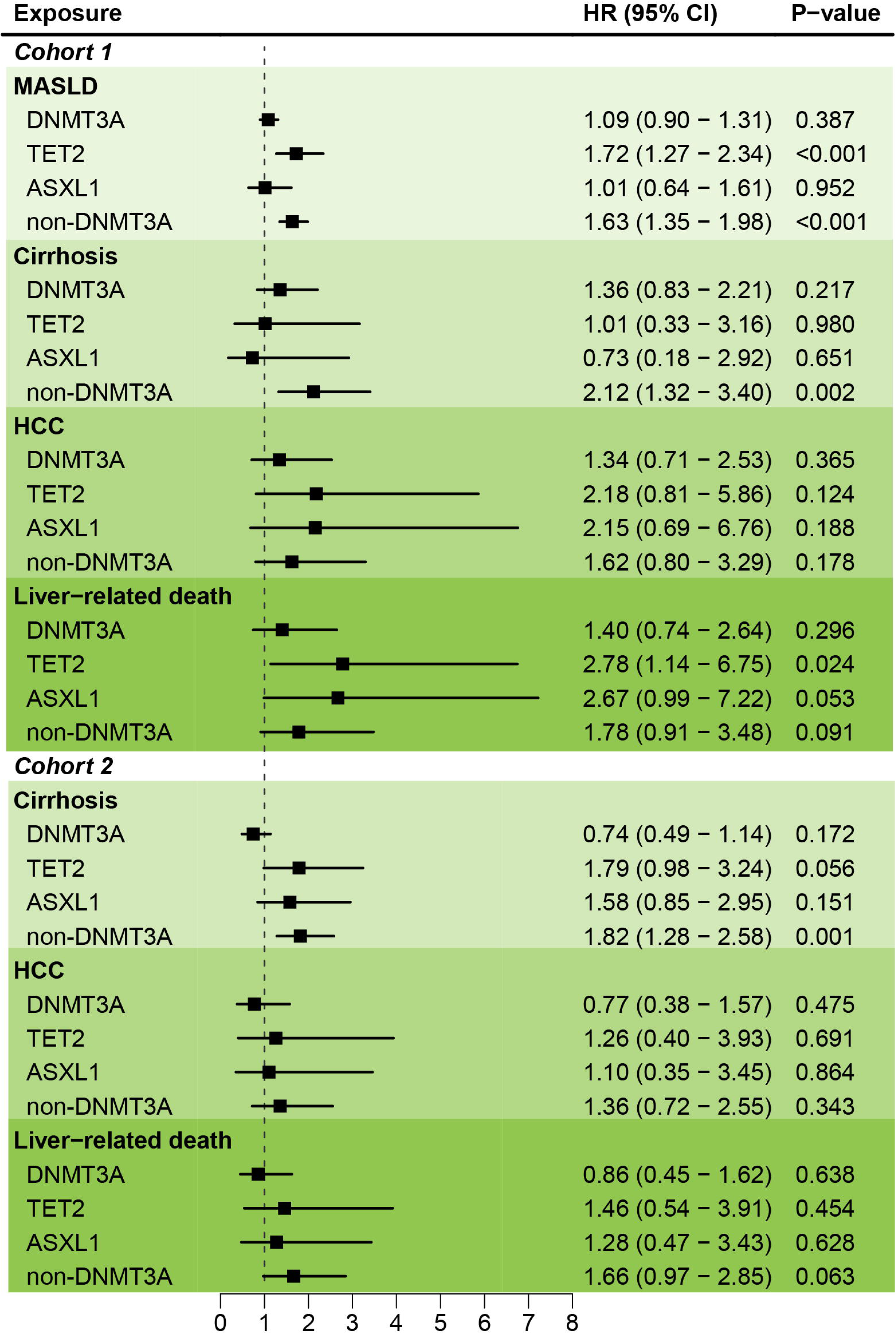
Gene-Specific Associations of CHIP with Liver Outcomes in Both Cohorts Forest plot showing HRs and 95% CIs for the associations of CHIP, stratified by driver gene (DNMT3A, TET2, ASXL1, and non-DNMT3A), with liver-related outcomes in Cohort 1 (incident MASLD, cirrhosis, HCC, and liver-related death) and Cohort 2 (cirrhosis, HCC, and liver-related death). Results are from the fully adjusted Cox proportional hazards model (Model 3), adjusted for age, sex, ethnicity, education, Townsend deprivation index, smoking status, physical activity, daily alcohol intake, diet risk score, BMI, prevalent diabetes, hypertension, statin use, and baseline levels of triglycerides, HDL cholesterol, CRP, ALT, GGT, and platelet count. **Abbreviations:** CHIP, clonal hematopoiesis of indeterminate potential; CI, confidence interval; HCC, hepatocellular carcinoma; HR, hazard ratio; MASLD, metabolic dysfunction-associated steatotic liver disease.

**Figure 4.**
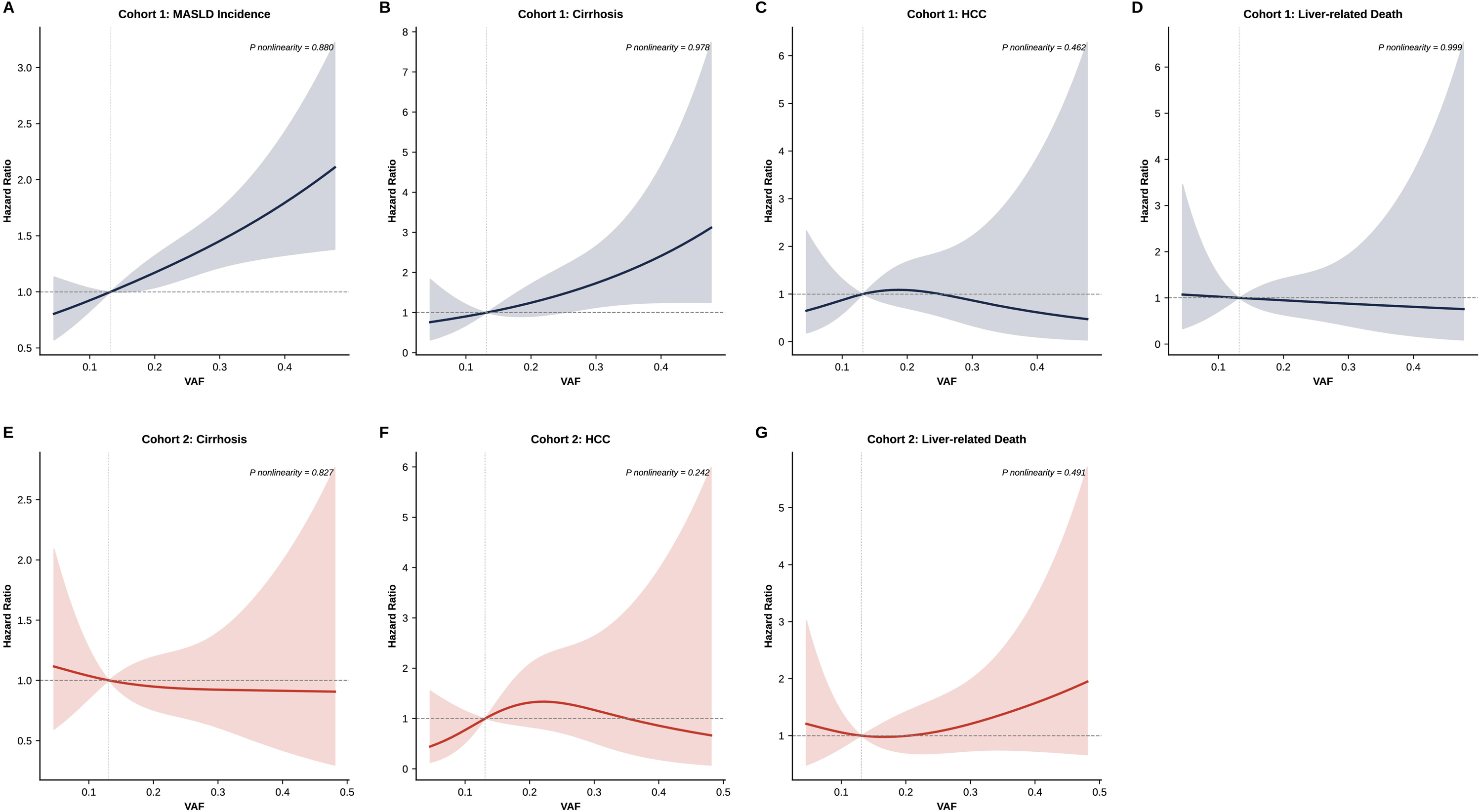
Dose-Response Relationship Between CHIP Clone Size and Liver Outcomes Restricted cubic spline plots illustrating the dose-response relationship between CHIP VAF and the risk of liver-related outcomes in Cohort 1 (Panel A: incident MASLD; Panel B: incident cirrhosis) and Cohort 2 (Panel C: cirrhosis; Panel D: liver-related death). The solid lines represent the estimated hazard ratios, and the shaded areas represent the 95% confidence intervals. Splines were modeled with four knots. P-values for non-linearity are shown. Models were fully adjusted as described in Figure 3. **Abbreviations:** CI, confidence interval; HR, hazard ratio; MASLD, metabolic dysfunction-associated steatotic liver disease; VAF, variant allele fraction.

### CHIP and risk of liver disease progression in the MASLD population

In Cohort 2, with a median follow-up of 12.8 years, we observed 1,150 incident cirrhosis cases, 356 incident HCC cases, and 412 liver-related deaths (**Figure 2**). Gene-specific analyses revealed that non-DNMT3A mutations were significantly associated with an 82% increased risk of progression to cirrhosis (HR 1.82, 95% CI 1.28-2.58), while TET2 mutations showed a borderline association (HR 1.79, 95% CI 0.98-3.24; P=0.056) (**Figure 3**). Non-DNMT3A mutations also showed a trend toward increased liver-related death (HR 1.66, 95% CI 0.97-2.85; P=0.063). In contrast to Cohort 1, small CHIP was associated with a 61% increased risk of progression to cirrhosis (HR 1.61, 95% CI 1.03-2.51). No significant dose-response relationships were observed in this cohort (P for non-linearity >0.05 for all outcomes) (**Figure 4**). The main results were consistent in competing risk analyses (**Supplementary Figure S2 and S3**).

### Association of CHIP with hepatic and inflammatory biomarkers

In cross-sectional analyses, the presence of CHIP was associated with a pro-inflammatory and pro-fibrotic biomarker profile (**Supplementary Table S5**). Specifically, TET2-mutant CHIP was associated with higher odds of low HDL cholesterol (OR 1.30, 95% CI 1.13-1.50) and elevated FIB-4 (OR 1.16, 95% CI 1.02-1.32). Non-DNMT3A CHIP was associated with elevated CRP (OR 1.13, 95% CI 1.04-1.23), elevated FIB-4 (OR 1.11, 95% CI 1.01-1.22), and low HDL cholesterol (OR 1.29, 95% CI 1.18-1.40). In contrast, DNMT3A-mutant CHIP was associated with elevated blood pressure (OR 1.16, 95% CI 1.08-1.24) but lower odds of having an elevated FIB-4 score (OR 0.90, 95% CI 0.84-0.97).

### Subgroup analyses

The associations between the presence of CHIP and liver outcomes were largely consistent across pre-specified subgroups of age, sex, BMI, smoking status, and ethnicity, with most P-values for interaction being >0.05 (**Figure 5**). In Cohort 1, the increased risk of MASLD associated with CHIP was particularly evident in older individuals (age ≥60 years: HR 1.32, 95% CI 1.12-1.55) and males (HR 1.38, 95% CI 1.10-1.73). In Cohort 2, we observed significant effect modification by sex (P-interaction=0.045), BMI (P-interaction<0.001), and diabetes status (P-interaction=0.023). The association of CHIP with liver disease progression was more pronounced in males (HR 1.49, 95% CI 1.05-2.12) than in females (HR 0.87, 95% CI 0.55-1.39), in participants with normal weight (HR 2.34, 95% CI 0.81-6.78) compared to those with obesity (HR 0.58, 95% CI 0.38-0.89), and in those without diabetes (HR 1.44, 95% CI 1.04-2.00) compared to those with diabetes (HR 0.74, 95% CI 0.44-1.27).

**Figure 5.**
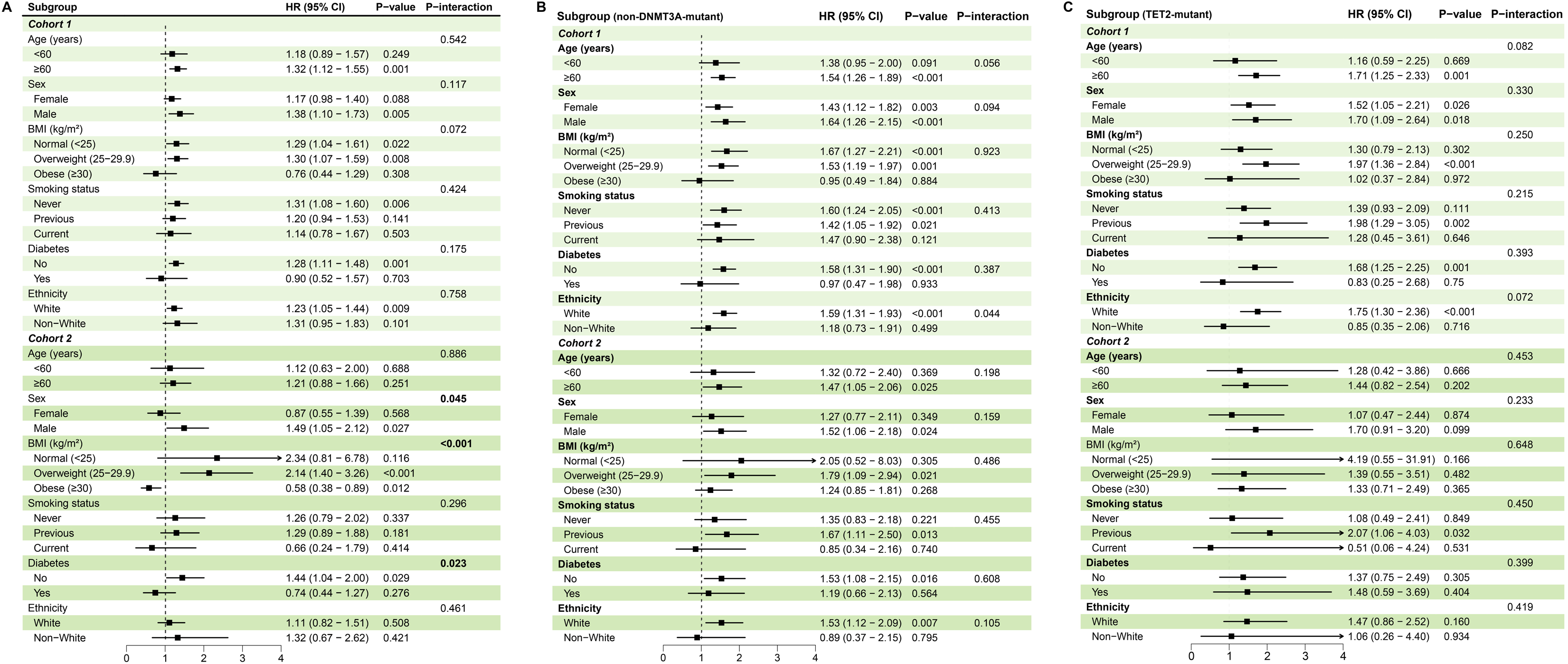
Subgroup Analyses of the Association Between CHIP and Liver Outcomes Forest plot showing HRs and 95% CIs for the association between any CHIP and liver outcomes across pre-specified subgroups of age, sex, BMI, smoking status, prevalent diabetes, and ethnicity. Results are shown for incident MASLD in Cohort 1 and cirrhosis in Cohort 2. P-values for interaction were derived from multiplicative interaction terms in the fully adjusted Cox model. **Abbreviations:** BMI, body mass index; CHIP, clonal hematopoiesis of indeterminate potential; CI, confidence interval; HR, hazard ratio; MASLD, metabolic dysfunction-associated steatotic liver disease.

### Mediation analyses

In mediation analyses, inflammatory biomarkers partially explained the association between CHIP and liver outcomes (**Figure 6**). In Cohort 1, CRP mediated 2.2% (P=0.014) of the total effect of CHIP on incident MASLD and 1.2% (P=0.026) of the effect on incident cirrhosis. White blood cell count and the systemic immune-inflammation index (SII) also showed significant mediating effects for MASLD (1.7% and 1.1%, respectively). In Cohort 2, monocyte count mediated 2.2% (P=0.019) of the association with cirrhosis progression, while AST appeared to be a strong mediator (13.1%), although this did not reach statistical significance (P=0.092). Among 25 tested cytokines, most showed no significant mediating effect, with the exception of IL-10, which mediated a small proportion of the association between CHIP and liver-related death in Cohort 2 (**Supplementary Figure S4**).

**Figure 6.**
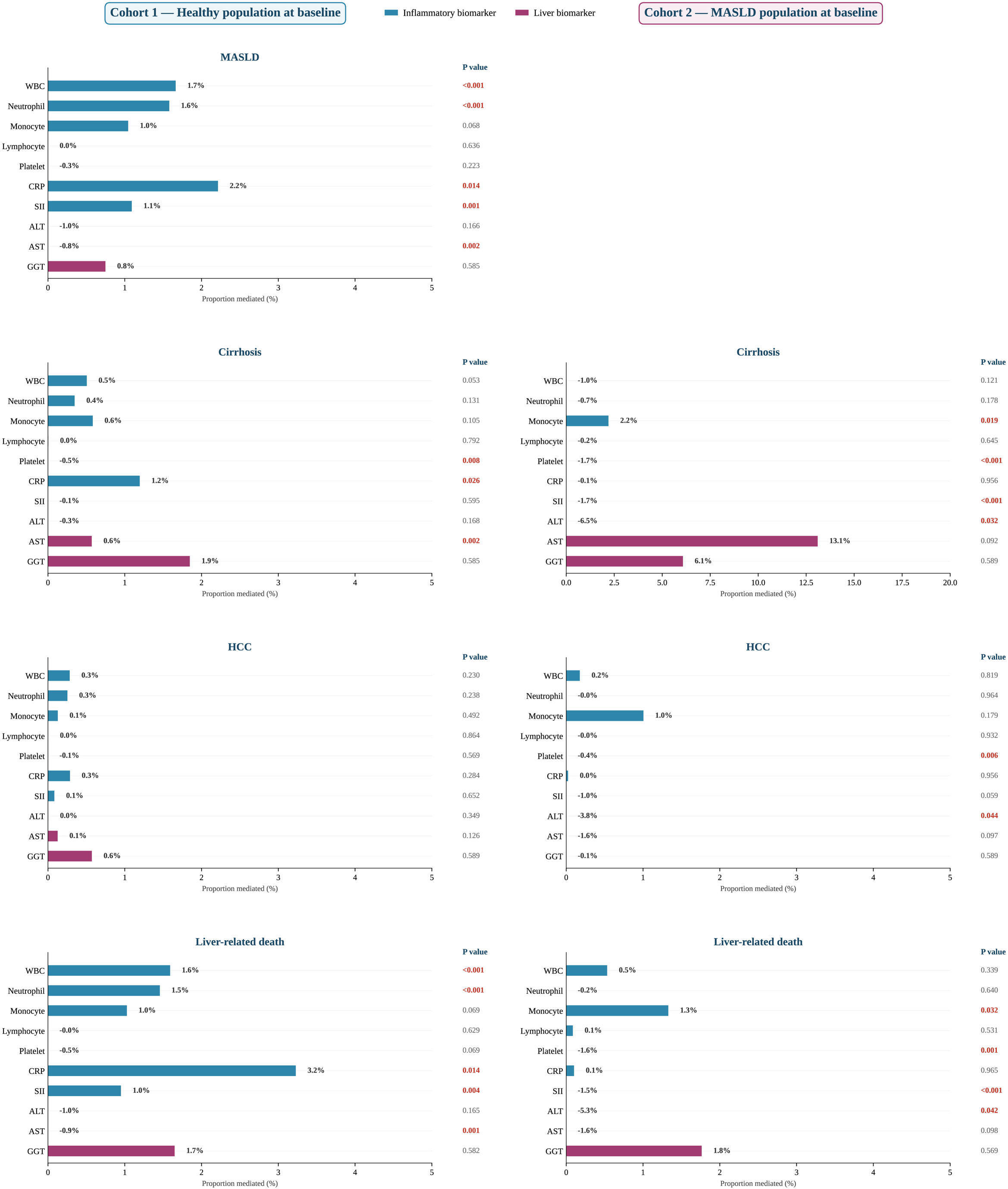
Mediation of the Association Between CHIP and Liver Outcomes by Inflammatory Biomarkers Bar plot showing the proportion of the total effect of CHIP on liver outcomes mediated by candidate inflammatory and hepatic biomarkers, including white blood cell count, neutrophil count, monocyte count, lymphocyte count, platelet count, C-reactive protein (CRP), systemic immune-inflammation index (SII), alanine aminotransferase (ALT), aspartate aminotransferase (AST), and gamma-glutamyl transferase (GGT). Results are shown for Cohort 1 (incident MASLD and cirrhosis) and Cohort 2 (cirrhosis and liver-related death). The proportion mediated and 95% confidence intervals were estimated using a causal mediation framework with 1,000 bootstrap resamples. Asterisks indicate statistical significance (*P<0.05, **P<0.01). **Abbreviations:** ALT, alanine aminotransferase; AST, aspartate aminotransferase; CHIP, clonal hematopoiesis of indeterminate potential; CRP, C-reactive protein; GGT, gamma-glutamyl transferase; MASLD, metabolic dysfunction-associated steatotic liver disease; SII, systemic immune-inflammation index.

## Discussion

In this large-scale, prospective study of over 350,000 individuals from the UK Biobank, we provide comprehensive evidence that CHIP is associated with both the initial development of MASLD and its progression to severe clinical endpoints, including cirrhosis and liver-related death. Our findings demonstrate that this risk is primarily driven by mutations in non-DNMT3A genes, particularly TET2, and exhibits a dose-dependent relationship with clone size. Furthermore, we identify distinct patterns of association between specific CHIP driver genes and cardiometabolic biomarkers, and provide evidence that systemic inflammation partially mediates the observed associations.

Our results substantially extend the evidence of recent landmark studies. A pivotal 2023 study by Wong et al. was the first to demonstrate a robust association between CHIP and the progression of chronic liver disease (CLD) in a pooled analysis of four cohorts (N≈215,000).^[15]^ Our findings in the MASLD progression cohort (Cohort 2) are consistent with theirs, showing that the presence of CHIP, particularly non-DNMT3A mutations, is associated with an increased risk of cirrhosis. However, our study provides a critical new dimension by establishing CHIP as a risk factor for incident MASLD in a large, initially healthy population (Cohort 1), a distinction not made in prior research. This suggests that CHIP may not only accelerate the progression of established liver disease but may also contribute to its initial development. Our findings are also in line with a recent cross-sectional study by Marchetti et al. (N=640), which reported an association between CHIP (especially TET2) and HCC in individuals with steatotic liver disease.^[16]^ Although our association between TET2 and incident HCC did not reach statistical significance, the elevated point estimate (HR 2.18) suggests a consistent trend that may have been limited by the low number of incident HCC cases. Importantly, our detailed gene-specific analyses reveal that the risk is not uniformly distributed across CHIP driver genes. While non-DNMT3A and TET2 mutations consistently showed elevated risks, DNMT3A-mutant CHIP—the most common CHIP subtype—was not significantly associated with any liver endpoint.^[6]^ This distinction, which has been hinted at in cardiovascular literature, has not been clearly delineated in the context of liver disease and has important implications for risk stratification.^[10, 27, 28]^

The differential effects of CHIP driver genes observed in our study point towards distinct underlying biological mechanisms. The strong association of non-DNMT3A mutations (predominantly TET2) with liver outcomes and inflammatory biomarkers like CRP supports the prevailing hypothesis that CHIP exerts its pathogenic effects through a pro-inflammatory state.

Mechanistic studies have shown that loss of TET2 function in macrophages leads to hyperactivation of the NLRP3 inflammasome and increased production of pro-inflammatory cytokines like IL-1β and IL-6, which are known drivers of liver inflammation and fibrosis.^[11, 12]^ Our mediation analysis, showing that CRP partially explains the association between CHIP and incident MASLD, provides human epidemiological evidence for this inflammatory pathway. However, the mediation proportion was modest (2.2%), suggesting that additional mechanisms are involved. Our broader cytokine mediation analysis using the Olink proteomics platform was largely null, with only IL-10 showing a significant, albeit small, mediation effect for liver-related death in individuals with MASLD. This general lack of significant mediation by circulating cytokines may reflect several factors: the limited sample size of the Olink sub-cohort, the possibility that single time-point measurements in peripheral blood inadequately capture chronic, low-grade inflammation within the hepatic microenvironment, or the involvement of unmeasured inflammatory pathways.^[29]^ In contrast to TET2, DNMT3A-mutant CHIP showed no significant association with liver outcomes. This aligns with findings from cardiovascular research, where DNMT3A and TET2 mutations have also been shown to have divergent effects on atherosclerosis and heart failure. A recent mouse model study by Yan et al. did report that Dnmt3a-driven CH promoted MASLD and fibrosis;^[30]^ however, this was in a model with a large (20%) clonal fraction, which may not be representative of the smaller clone sizes often seen in human DNMT3A CHIP. The association we observed between DNMT3A mutations and elevated blood pressure, but a lower risk of elevated FIB-4, further suggests that DNMT3A may influence systemic metabolic health through non-inflammatory or even counter-regulatory mechanisms that warrant further investigation.^[31]^

An intriguing finding of our study was the significant modification of the CHIP-liver disease association by sex, BMI, and diabetes status in the MASLD progression cohort. The stronger association observed in males aligns with established sex differences in MASLD, where premenopausal women are relatively protected, likely due to the hepatoprotective effects of estrogen, which can suppress hepatic inflammation.^[32, 33]^ The attenuated or even inverted association in individuals with obesity or diabetes is also noteworthy. We hypothesize that in these individuals, the substantial pre-existing inflammatory and metabolic burden driven by obesity or diabetes may attenuate the incremental pathogenic contribution of CHIP, as the baseline level of hepatic inflammation is already sufficiently high to drive disease progression, thereby diminishing the relative impact of CHIP-mediated inflammation.^[34]^ Conversely, in non-obese, non-diabetic individuals, the pro-inflammatory state induced by CHIP may represent a more significant pathogenic driver. These observations suggest that the clinical impact of CHIP on liver disease progression may be most pronounced in individuals without other major metabolic comorbidities and highlight the complex interplay between genetic predisposition and metabolic health.

Our findings have important clinical implications. Given the high prevalence of both MASLD and CHIP in the aging population, our results identify CHIP as a novel risk factor for the entire spectrum of steatotic liver disease, with potential implications for targeted therapeutic intervention.^[2, 35]^ This raises the possibility that individuals with CHIP, particularly those with non-DNMT3A mutations, could be candidates for more intensive liver surveillance or targeted preventive strategies. The observation that even small clones (VAF <10%) were associated with an increased risk of cirrhosis progression in the MASLD cohort suggests that the risk is not confined to those with large clonal expansions. Furthermore, the partial mediation by CRP suggests that anti-inflammatory therapies, such as NLRP3 inhibitors currently in development for NASH, could be a promising therapeutic avenue for mitigating the liver-related risks associated with CHIP.^[36]^ The CANTOS trial, which showed that IL-1β inhibition reduced cardiovascular events in individuals with elevated CRP, provides a proof-of-concept for this strategy.^[37]^ Whether such therapies would be particularly beneficial for individuals with CHIP-associated liver disease warrants investigation in future clinical trials.

This study has several major strengths, including its large sample size, prospective design, and the availability of centrally adjudicated, genome-wide CHIP data. By analyzing two distinct cohorts, we were able to separately assess the role of CHIP in both the incidence and progression of MASLD, a key distinction not made in prior research. The comprehensive assessment of multiple liver-related outcomes, detailed gene- and clone size-specific analyses, and the inclusion of mediation and biomarker analyses provide a granular view of the CHIP-liver disease relationship. However, our study also has limitations. First, as an observational study, we cannot definitively establish causality, although the consistency of our findings with experimental data and the linear dose-response relationship provide supportive evidence. Second, the diagnosis of MASLD was based on the FLI rather than imaging, which may lead to some misclassification, although FLI is a widely used and validated tool for epidemiological studies.^[18, 20]^ Third, the UK Biobank population is predominantly of European ancestry, which may limit the generalizability of our findings to other ethnic groups.^[38]^ Finally, despite the large sample size, the number of incident events for some outcomes, such as HCC, was low, which may have limited our statistical power to detect associations for less frequent CHIP mutations.

In conclusion, our findings from this large, prospective study demonstrate that CHIP is a significant risk factor for both the development and progression of MASLD. The risk is primarily driven by non-DNMT3A mutations, is dose-dependent on clone size, and appears to be at least partially mediated by systemic inflammation. These findings position CHIP as an important player in the pathophysiology of MASLD and open new avenues for risk stratification and therapeutic intervention in this increasingly common liver disease.

## Abbreviations

ALT: Alanine aminotransferase
AST: Aspartate aminotransferase
BMI: Body mass index
CHIP: Clonal hematopoiesis of indeterminate potential
CI: Confidence interval
CRP: C-reactive protein
FIB-4: Fibrosis-4 index
FLI: Fatty Liver Index
GATK: Genome Analysis Toolkit
GGT: Gamma-glutamyl transferase
HCC: Hepatocellular carcinoma
HDL: High-density lipoprotein
HES: Hospital Episode Statistics
HR: Hazard ratio
ICD-10: International Classification of Diseases, 10th Revision
IL: Interleukin
IQR: Interquartile range
MASLD: Metabolic dysfunction-associated steatotic liver disease
NAFLD: Non-alcoholic fatty liver disease
NASH: Non-alcoholic steatohepatitis
NLRP3: NLR family pyrin domain containing 3
OR: Odds ratio
PEDW: Patient Episode Database for Wales
SD: Standard deviation
SHR: Subdistribution hazard ratio
SII: Systemic immune-inflammation index
SMR: Scottish Morbidity Records
VAF: Variant allele fraction
WES: Whole-exome sequencing

## Acknowledgements

This research was conducted using the UK Biobank Resource under Application Number 101633. We thank all participants of the UK Biobank and the staff of the UK Biobank assessment centres for their contributions to this resource.

## Authors’ contributions

Ruijie Xie: Conceptualization, Methodology, Formal analysis, Data curation, Writing - original draft, Visualization. Ben Schöttker: Conceptualization, Methodology, Supervision, Writing - review & editing, Project administration. All authors read and approved the final manuscript. Ruijie Xie and Ben Schöttker are the guarantors of this work and, as such, had full access to all the data in the study and take responsibility for the integrity of the data and the accuracy of the data analysis.

## Data availability statement

Data from the UK Biobank are available to bona fide researchers upon application through the UK Biobank Access Management System (https://www.ukbiobank.ac.uk/enable-your-research/apply-for-access). The analytical code supporting the findings of this study is available from the corresponding author upon reasonable request.

---

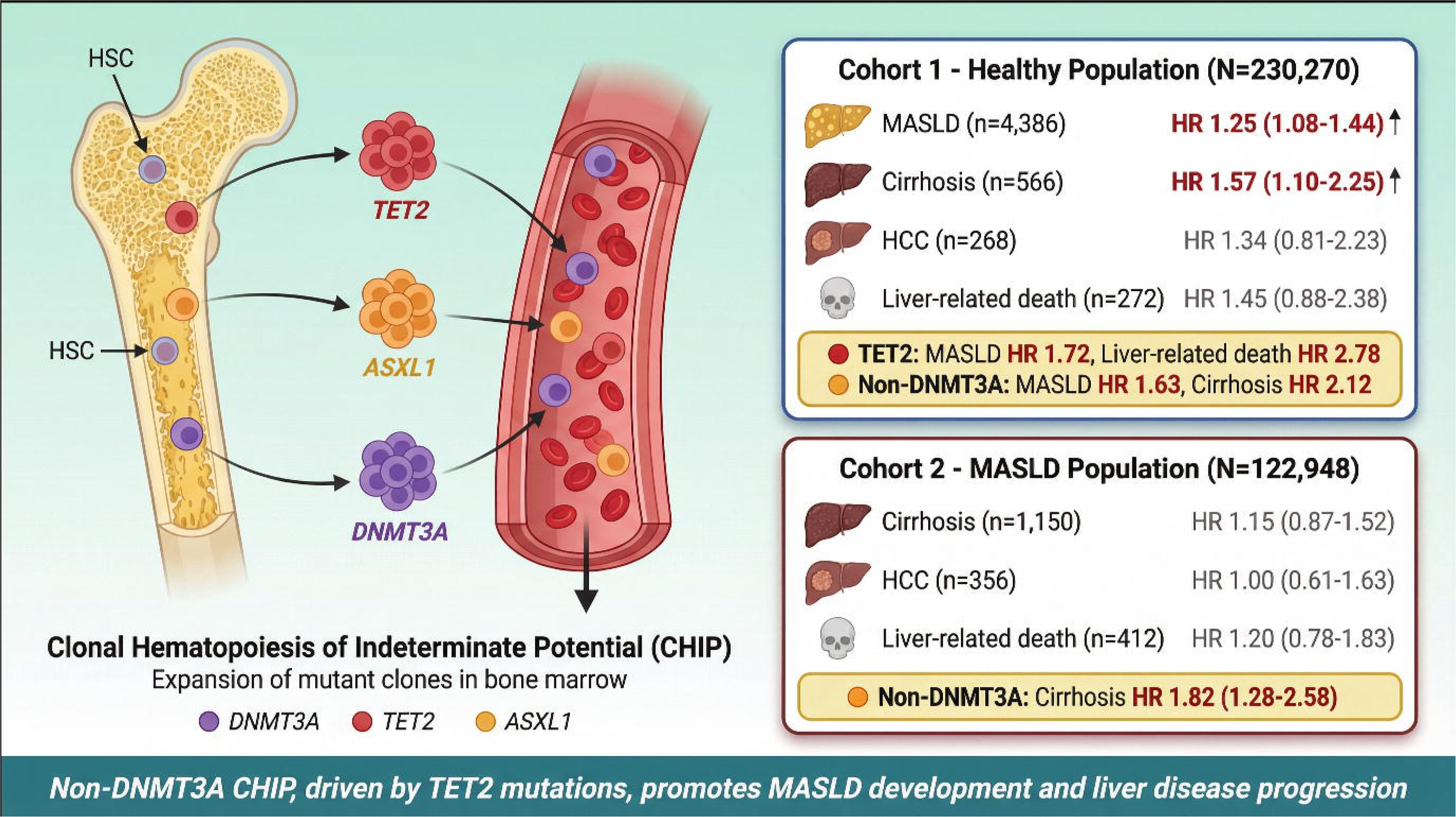

## References

[1] Rinella ME, Lazarus JV, Ratziu V, et al. A multisociety Delphi consensus statement on new fatty liver disease nomenclature. Hepatology. 2023;78:1966–1986.

[2] Younossi ZM, Golabi P, Paik JM, et al. The global epidemiology of nonalcoholic fatty liver disease (NAFLD) and nonalcoholic steatohepatitis (NASH): a systematic review. Hepatology. 2023;77:1335–1347.

[3] Powell EE, Wong VW-S, Rinella M. Non-alcoholic fatty liver disease. The Lancet. 2021;397:2212–2224.

[4] Huang DQ, El-Serag HB, Loomba R. Global epidemiology of NAFLD-related HCC: trends, predictions, risk factors and prevention. Nature reviews Gastroenterology & hepatology. 2021;18:223–238.

[5] Taylor RS, Taylor RJ, Bayliss S, et al. Association between fibrosis stage and outcomes of patients with nonalcoholic fatty liver disease: a systematic review and meta-analysis. Gastroenterology. 2020;158:1611–1625. e1612.

[6] Steensma DP, Bejar R, Jaiswal S, et al. Clonal hematopoiesis of indeterminate potential and its distinction from myelodysplastic syndromes. Blood, The Journal of the American Society of Hematology. 2015;126:9–16.

[7] Marnell CS, Bick A, Natarajan P. Clonal hematopoiesis of indeterminate potential (CHIP): Linking somatic mutations, hematopoiesis, chronic inflammation and cardiovascular disease. J Mol Cell Cardiol. 2021;161:98–105.

[8] Jaiswal S, Fontanillas P, Flannick J, et al. Age-related clonal hematopoiesis associated with adverse outcomes. N Engl J Med. 2014;371:2488–2498.

[9] Genovese G, Kähler AK, Handsaker RE, et al. Clonal hematopoiesis and blood-cancer risk inferred from blood DNA sequence. N Engl J Med. 2014;371:2477–2487.

[10] Jaiswal S, Natarajan P, Silver AJ, et al. Clonal hematopoiesis and risk of atherosclerotic cardiovascular disease. N Engl J Med. 2017;377:111–121.

[11] Fuster JJ, MacLauchlan S, Zuriaga MA, et al. Clonal hematopoiesis associated with TET2 deficiency accelerates atherosclerosis development in mice. Science. 2017;355:842–847.

[12] Sano S, Oshima K, Wang Y, et al. Tet2-mediated clonal hematopoiesis accelerates heart failure through a mechanism involving the IL-1β/NLRP3 inflammasome. J Am Coll Cardiol. 2018;71:875–886.

[13] Kazankov K, Jørgensen SMD, Thomsen KL, et al. The role of macrophages in nonalcoholic fatty liver disease and nonalcoholic steatohepatitis. Nature reviews Gastroenterology & hepatology. 2019;16:145–159.

[14] Tacke F. Targeting hepatic macrophages to treat liver diseases. J Hepatol. 2017;66:1300–1312.

[15] Wong WJ, Emdin C, Bick AG, et al. Clonal haematopoiesis and risk of chronic liver disease. Nature. 2023;616:747–754.

[16] Marchetti A, Pelusi S, Marella A, et al. Impact of clonal hematopoiesis of indeterminate potential on hepatocellular carcinoma in individuals with steatotic liver disease. Hepatology. 2024;80:816–827.

[17] Sudlow C, Gallacher J, Allen N, et al. UK biobank: an open access resource for identifying the causes of a wide range of complex diseases of middle and old age. PLoS Med. 2015;12:e1001779.

[18] Bedogni G, Bellentani S, Miglioli L, et al. The Fatty Liver Index: a simple and accurate predictor of hepatic steatosis in the general population. BMC Gastroenterol. 2006;6:33.

[19] Feng Q, Izzi-Engbeaya CN, Branch AD, et al. The Relationships Between MASLD, Extrahepatic Multimorbidity, and All-Cause Mortality in the UK Biobank Cohort. The Journal of Clinical Endocrinology & Metabolism. 2026;111:e58–e69.

[20] Carli F, Sabatini S, Gaggini M, et al. Fatty liver index (FLI) identifies not only individuals with liver steatosis but also at high cardiometabolic risk. Int J Mol Sci. 2023;24:14651.

[21] Vlasschaert C, Mack T, Heimlich JB, et al. A practical approach to curate clonal hematopoiesis of indeterminate potential in human genetic data sets. Blood, The Journal of the American Society of Hematology. 2023;141:2214–2223.

[22] Hu B, Yang XR, Xu Y, et al. Systemic immune-inflammation index predicts prognosis of patients after curative resection for hepatocellular carcinoma. Clin Cancer Res. 2014;20:6212–6222.

[23] Sterling RK, Lissen E, Clumeck N, et al. Development of a simple noninvasive index to predict significant fibrosis in patients with HIV/HCV coinfection. Hepatology. 2006;43:1317–1325.

[24] Stekhoven DJ, Bühlmann P. MissForest—non-parametric missing value imputation for mixed-type data. Bioinformatics. 2012;28:112–118.

[25] Fine JP, Gray RJ. A proportional hazards model for the subdistribution of a competing risk. Journal of the American statistical association. 1999;94:496–509.

[26] VanderWeele TJ. Mediation analysis: a practitioner’s guide. Annu Rev Public Health. 2016;37:17–32.

[27] Dorsheimer L, Assmus B, Rasper T, et al. Association of mutations contributing to clonal hematopoiesis with prognosis in chronic ischemic heart failure. JAMA Cardiol. 2019;4:25–33.

[28] Jaiswal S, Libby P. Clonal haematopoiesis: connecting ageing and inflammation in cardiovascular disease. Nature Reviews Cardiology. 2020;17:137–144.

[29] Libby P. Interleukin-1 beta as a target for atherosclerosis therapy: biological basis of CANTOS and beyond. J Am Coll Cardiol. 2017;70:2278–2289.

[30] Yan B, Yuan Q, Buttigieg MM, et al. Clonal hematopoiesis driven by Dnmt3a mutations promotes metabolic disease development in mice. The Journal of Clinical Investigation. 2025;135.

[31] Bick AG, Weinstock JS, Nandakumar SK, et al. Inherited causes of clonal haematopoiesis in 97,691 whole genomes. Nature. 2020;586:763–768.

[32] Lonardo A, Nascimbeni F, Ballestri S, et al. Sex differences in nonalcoholic fatty liver disease: state of the art and identification of research gaps. Hepatology. 2019;70:1457–1469.

[33] Ezhilarasan D. Critical role of estrogen in the progression of chronic liver diseases. Hepatobiliary Pancreat Dis Int. 2020;19:429–434.

[34] Henn RE, Elzinga SE, Glass E, et al. Obesity-induced neuroinflammation and cognitive impairment in young adult versus middle-aged mice. Immun Ageing. 2022;19:67.

[35] Steensma DP. Clinical consequences of clonal hematopoiesis of indeterminate potential. Blood Adv. 2018;2:3404–3410.

[36] Mridha AR, Wree A, Robertson AA, et al. NLRP3 inflammasome blockade reduces liver inflammation and fibrosis in experimental NASH in mice. J Hepatol. 2017;66:1037–1046.

[37] Ridker PM, Everett BM, Thuren T, et al. Antiinflammatory Therapy with Canakinumab for Atherosclerotic Disease. N Engl J Med. 2017;377:1119–1131.

[38] Bycroft C, Freeman C, Petkova D, et al. The UK Biobank resource with deep phenotyping and genomic data. Nature. 2018;562:203–209.

