## Supplementary Material for "Clonal Hematopoiesis and the Development and Progression of Metabolic Dysfunction-Associated Steatotic Liver Disease"

**Supplementary Table S1.** Definitions of Exclusion Criteria

| **Exclusion Criterion** | **Definition** | **ICD-10 Codes / Criteria** | **N Excluded** |
| --- | --- | --- | --- |
| **Heavy Alcohol Consumption** | Average daily alcohol intake >30 g for men or >20 g for women. | — | 106,551 |
| **Viral Hepatitis** | Prevalent diagnosis of chronic viral hepatitis at or before baseline. | B18 | 538 |
| **Toxic Liver Disease** | Prevalent diagnosis of toxic liver disease at or before baseline. | K71 | 52 |
| **Advanced Liver Disease** | Prevalent diagnosis of hepatic fibrosis, cirrhosis, or alcoholic cirrhosis at or before baseline. | K74, K70.3 | 974 |
| **Unavailable CHIP Data** | Participants without CHIP return data in the UK Biobank dataset. | — | 40,603 |

Note: Exclusions were applied sequentially.

**Supplementary Table S2.** Definitions of Cardiometabolic Risk Factors for MASLD Ascertainment

| **Risk Factor** | **Definition** | **Criteria / ICD-10 Codes** |
| --- | --- | --- |
| **Overweight / Obesity** | Body mass index (BMI) ≥25 kg/m². | BMI ≥25 kg/m² |
| **Type 2 Diabetes** | Physician diagnosis, use of glucose-lowering medication, or HbA_1c_ ≥48 mmol/mol (6.5%). | E11; or HbA_1c_ ≥48 mmol/mol; or medication |
| **Hypertension** | Systolic blood pressure ≥130 mmHg, diastolic blood pressure ≥85 mmHg, or use of antihypertensive medication. | SBP ≥130 or DBP ≥85 mmHg; or I10–I15; or medication |
| **Hypertriglyceridemia** | Fasting serum triglycerides ≥1.7 mmol/L. | Triglycerides ≥1.7 mmol/L |
| **Low HDL Cholesterol** | Fasting serum HDL cholesterol <1.03 mmol/L for men or <1.29 mmol/L for women. | HDL <1.03 mmol/L (men) or <1.29 mmol/L (women) |

Note: MASLD was defined as hepatic steatosis (FLI ≥60) plus at least one of the above cardiometabolic risk factors.

**Supplementary Table S3.** Definitions of Clinical Outcomes

| **Outcome** | **Definition** | **ICD-10 Codes** | **Data Source** |
| --- | --- | --- | --- |
| **Incident MASLD** | First occurrence of a MASLD-related diagnosis code after baseline, in participants free of MASLD at baseline. | K76.0, K75.8 | Hospital Inpatient, Primary Care |
| **Incident Cirrhosis** | First occurrence of a cirrhosis-related diagnosis code after baseline. | K74.0-K74.6 | Hospital Inpatient, Primary Care, Death Registry |
| **Incident HCC** | First occurrence of a hepatocellular carcinoma diagnosis code after baseline. | C22.0 | Cancer Registry, Death Registry |
| **Liver-Related Death** | Death with a liver condition listed as the primary or contributory cause of death. | K70–K77, C22.0 | National Death Registry |

**Abbreviations:** MASLD, metabolic dysfunction-associated steatotic liver disease; HCC, hepatocellular carcinoma; ICD-10, International Classification of Diseases, 10th Revision.

**Supplementary Table S4.** Definitions and Measurement of Covariates

| **Category** | **Covariate** | **Definition** | **Categories** | **Measurement Method** |
| --- | --- | --- | --- | --- |
| **Sociodemographic** | Age | Age at baseline assessment | Continuous (years) | Self-report |
| **Sociodemographic** | Sex | Genetic sex | Male / Female | Self-report |
| **Sociodemographic** | Ethnicity | Self-reported ethnic background | White / Non-White | Self-report Questionnaire |
| **Sociodemographic** | Education Level | Highest educational qualification | High / Intermediate / Low | Self-report Questionnaire |
| **Sociodemographic** | Townsend Deprivation Index | Area-level socioeconomic deprivation score | Continuous | Geocoded Linkage |
| **Lifestyle** | Smoking Status | Self-reported smoking behavior | Never / Previous / Current | Self-report Questionnaire |
| **Lifestyle** | Physical Activity | Level of physical activity | Low / Moderate / High (IPAQ) | Self-report Questionnaire |
| **Lifestyle** | Daily Alcohol Intake | Grams of alcohol consumed per day | Continuous (g/day) | Self-report Questionnaire |
| **Lifestyle** | Diet Risk Score | Composite score of dietary habits | Continuous | Self-report Questionnaire |
| **Lifestyle** | Body Mass Index (BMI) | Weight in kilograms divided by height in meters squared | Continuous (kg/m²) | Tanita BC-418 MA / Seca 202 |
| **Clinical** | Diabetes | Prevalent Type 2 diabetes at baseline | Yes / No | Self-report, Medication, HbA1c |
| **Clinical** | Hypertension | Prevalent hypertension at baseline | Yes / No | Self-report, Medication, BP measurement |
| **Clinical** | Statin Use | Use of cholesterol-lowering statin medication | Yes / No | Self-report, Medication Records |
| **Biomarker** | Triglycerides | Serum triglycerides | Continuous (mmol/L) | Beckman Coulter AU5800 (Enzymatic) |
| **Biomarker** | HDL Cholesterol | Serum high-density lipoprotein cholesterol | Continuous (mmol/L) | Beckman Coulter AU5800 (Enzyme Immuno-inhibition) |
| **Biomarker** | C-reactive Protein (CRP) | High-sensitivity C-reactive protein | Continuous (mg/L, log-transformed) | Beckman Coulter AU5800 (Immuno-turbidimetric) |
| **Biomarker** | Alanine Aminotransferase (ALT) | Serum ALT | Continuous (U/L, log-transformed) | Beckman Coulter AU5800 (Enzymatic Rate) |
| **Biomarker** | Gamma-glutamyl Transferase (GGT) | Serum GGT | Continuous (U/L, log-transformed) | Beckman Coulter AU5800 (Enzymatic Rate) |
| **Biomarker** | Platelet Count | Platelet count in whole blood | Continuous (×10⁹/L) | Beckman Coulter LH 750 |

**Abbreviations:** IPAQ, International Physical Activity Questionnaire; BMI, body mass index; HDL, high-density lipoprotein; CRP, C-reactive protein; ALT, alanine aminotransferase; GGT, gamma-glutamyl transferase.

**Supplementary Table S5. Association of CHIP with Liver and Metabolic Biomarkers in Total Population (N = 353,218)**

| **Outcome** | **N events /**  **N total** | **Any CHIP** | **DNMT3A** | **TET2** | **ASXL1** | **Non-DNMT3A** |
| --- | --- | --- | --- | --- | --- | --- |
|  |  | OR (95% CI)  P value | OR (95% CI)  P value | OR (95% CI)  P value | OR (95% CI)  P value | OR (95% CI)  P value |
| Elevated ALT (M>33/F>25 U/L) | 67,832 / 336,660 | **0.92 (0.87-0.98)**  **P = 0.006** | 0.96 (0.89-1.03)  P = 0.262 | **0.84 (0.72-0.98)**  **P = 0.026** | **0.72 (0.59-0.86)**  **P <0.001** | **0.86 (0.79-0.95)**  **P = 0.002** |
| Elevated AST (>40 U/L) | 14,303 / 335,586 | **0.85 (0.75-0.96)**  **P = 0.009** | 0.88 (0.76-1.03)  P = 0.105 | 0.83 (0.61-1.13)  P = 0.240 | 0.81 (0.57-1.13)  P = 0.215 | 0.84 (0.70-1.01)  P = 0.066 |
| Elevated GGT (M>50/F>32 U/L) | 73,917 / 336,617 | 0.97 (0.92-1.02)  P = 0.242 | 0.96 (0.90-1.03)  P = 0.308 | 0.98 (0.85-1.13)  P = 0.794 | **0.83 (0.70-0.99)**  **P = 0.033** | 0.98 (0.90-1.06)  P = 0.586 |
| Elevated CRP (>3 mg/L) | 76,554 / 336,060 | 1.05 (0.99-1.11)  P = 0.084 | 0.99 (0.92-1.06)  P = 0.777 | 1.03 (0.89-1.19)  P = 0.659 | 1.12 (0.96-1.32)  P = 0.149 | **1.13 (1.04-1.23)**  **P = 0.004** |
| FIB-4 ≥1.3 | 143,333 / 327,234 | **0.89 (0.85-0.94)**  **P <0.001** | **0.88 (0.83-0.94)**  **P <0.001** | **1.16 (1.02-1.32)**  **P = 0.023** | **0.84 (0.73-0.98)**  **P = 0.024** | 0.93 (0.86-1.00)  P = 0.058 |
| Hypertriglyceridemia (≥1.7 mmol/L) | 133,808 / 336,527 | 1.00 (0.95-1.05)  P = 0.958 | 1.02 (0.96-1.09)  P = 0.437 | 0.92 (0.81-1.04)  P = 0.173 | 0.96 (0.83-1.10)  P = 0.553 | 0.96 (0.89-1.03)  P = 0.275 |
| Low HDL (M<1.03/F<1.29 mmol/L) | 74,380 / 308,264 | **1.07 (1.01-1.13)**  **P = 0.028** | 0.94 (0.88-1.02)  P = 0.126 | **1.30 (1.13-1.50)**  **P <0.001** | **1.20 (1.02-1.42)**  **P = 0.026** | **1.29 (1.18-1.40)**  **P <0.001** |
| Elevated BP (SBP≥130/DBP≥85) | 22,6798 / 330,910 | **1.08 (1.02-1.14)**  **P = 0.006** | **1.16 (1.08-1.24)**  **P <0.001** | 1.00 (0.87-1.15)  P = 0.974 | 0.97 (0.82-1.14)  P = 0.683 | 0.96 (0.88-1.04)  P = 0.319 |
| Elevated Glucose (≥5.6 mmol/L) | 45,704 / 308,041 | 0.98 (0.92-1.05)  P = 0.596 | 0.98 (0.91-1.07)  P = 0.662 | 0.98 (0.84-1.15)  P = 0.827 | 0.96 (0.80-1.15)  P = 0.647 | 0.97 (0.88-1.07)  P = 0.560 |
| Central Obesity (M≥94/F≥80 cm) | 211,323 / 352,373 | 0.97 (0.91-1.04)  P = 0.362 | 0.95 (0.88-1.03)  P = 0.224 | 1.04 (0.88-1.23)  P = 0.640 | 1.02 (0.84-1.24)  P = 0.851 | 1.01 (0.91-1.12)  P = 0.875 |
| Metabolic Syndrome (≥3/5) | 115,312 / 353,212 | 1.02 (0.97-1.07)  P = 0.542 | 1.02 (0.96-1.09)  P = 0.550 | 1.04 (0.91-1.18)  P = 0.556 | 0.99 (0.86-1.15)  P = 0.894 | 1.02 (0.94-1.10)  P = 0.658 |

**Abbreviations:** ALT, alanine aminotransferase; AST, aspartate aminotransferase; GGT, gamma-glutamyl transferase; CRP, C-reactive protein; FIB-4, Fibrosis-4 index; HDL, high-density lipoprotein; BP, blood pressure; MetS, metabolic syndrome; OR, odds ratio; CI, confidence interval.

Cut-off values: ALT (M >33 / F >25 U/L, ACG 2017); AST (>40 U/L); GGT (M >50 / F >32 U/L); CRP (>3 mg/L, AHA/CDC); FIB-4 (≥1.3, AASLD/BSG); Triglycerides (≥1.7 mmol/L, ATP III); HDL (M <1.03 / F <1.29 mmol/L, ATP III); BP (SBP ≥130 / DBP ≥85 mmHg, ATP III); Glucose (≥5.6 mmol/L, ATP III); Waist (M ≥94 / F ≥80 cm, IDF); MetS (≥3/5 components, ATP III).

Models adjusted for age, sex, ethnicity, education, Townsend deprivation index, smoking, alcohol intake, BMI, and physical activity.

**Bold values** indicate statistical significance (P < 0.05).

**Supplementary Table S6.** Association Between CHIP and Liver Disease Outcomes: Cox Proportional Hazards Regression

| **Outcome** | **Exposure** | **N events** | **Model 1 HR (95% CI)** | **P-value** | **Model 2 HR (95% CI)** | **P-value** | **Model 3 HR (95% CI)** | **P-value** |
| --- | --- | --- | --- | --- | --- | --- | --- | --- |
| **Cohort 1: Healthy Population (N = 230,270)** | | | | | | | | |
| **MASLD** | Any CHIP | 4,386 | 1.27 (1.10–1.46) | <0.001 | 1.24 (1.08–1.43) | 0.003 | 1.25 (1.08–1.44) | 0.002 |
|  | Small CHIP |  | 0.99 (0.76–1.30) | 0.95 | 0.97 (0.74–1.27) | 0.85 | 0.98 (0.75–1.29) | 0.91 |
|  | Large CHIP |  | 1.41 (1.20–1.66) | <0.001 | 1.37 (1.17–1.62) | <0.001 | 1.38 (1.17–1.62) | <0.001 |
|  | **P for trend** |  |  | <0.001 |  | <0.001 |  | <0.001 |
| **Cirrhosis** | Any CHIP | 566 | 1.61 (1.13–2.31) | 0.009 | 1.51 (1.05–2.17) | 0.024 | 1.57 (1.10–2.25) | 0.014 |
|  | Small CHIP |  | 1.35 (0.70–2.62) | 0.37 | 1.30 (0.67–2.52) | 0.43 | 1.39 (0.72–2.69) | 0.33 |
|  | Large CHIP |  | 1.74 (1.15–2.65) | 0.009 | 1.61 (1.06–2.46) | 0.027 | 1.66 (1.09–2.52) | 0.019 |
|  | **P for trend** |  |  | 0.005 |  | 0.016 |  | 0.01 |
| **HCC** | Any CHIP | 268 | 1.37 (0.83–2.28) | 0.22 | 1.32 (0.80–2.20) | 0.28 | 1.34 (0.81–2.23) | 0.26 |
|  | Small CHIP |  | 1.83 (0.86–3.88) | 0.12 | 1.79 (0.84–3.79) | 0.13 | 1.83 (0.86–3.88) | 0.12 |
|  | Large CHIP |  | 1.15 (0.59–2.24) | 0.69 | 1.10 (0.56–2.14) | 0.78 | 1.11 (0.57–2.17) | 0.75 |
|  | **P for trend** |  |  | 0.43 |  | 0.49 |  | 0.43 |
| **Liver-related death** | Any CHIP | 272 | 1.48 (0.91–2.43) | 0.12 | 1.40 (0.86–2.30) | 0.18 | 1.45 (0.88–2.38) | 0.14 |
|  | Small CHIP |  | 2.16 (1.07–4.37) | 0.032 | 2.11 (1.04–4.27) | 0.039 | 2.19 (1.08–4.43) | 0.03 |
|  | Large CHIP |  | 1.16 (0.60–2.26) | 0.66 | 1.08 (0.55–2.11) | 0.82 | 1.11 (0.57–2.17) | 0.75 |
|  | **P for trend** |  |  | 0.33 |  | 0.40 |  | 0.33 |
| **Cohort 2: MASLD Population (N = 122,948)** | | | | | | | | |
| **Cirrhosis** | Any CHIP | 1,150 | 1.27 (0.97–1.68) | 0.088 | 1.24 (0.94–1.63) | 0.12 | 1.15 (0.87–1.52) | 0.32 |
|  | Small CHIP |  | 1.38 (0.89–2.15) | 0.15 | 1.35 (0.87–2.11) | 0.18 | 1.61 (1.03–2.51) | 0.035 |
|  | Large CHIP |  | 1.22 (0.86–1.71) | 0.26 | 1.18 (0.84–1.67) | 0.34 | 0.98 (0.69–1.39) | 0.91 |
|  | **P for trend** |  |  | 0.20 |  | 0.26 |  | 0.63 |
| **HCC** | Any CHIP | 356 | 1.08 (0.67–1.77) | 0.74 | 1.07 (0.66–1.74) | 0.79 | 1.00 (0.61–1.63) | 0.99 |
|  | Small CHIP |  | 0.76 (0.28–2.05) | 0.59 | 0.75 (0.28–2.02) | 0.57 | 0.84 (0.31–2.26) | 0.73 |
|  | Large CHIP |  | 1.25 (0.71–2.17) | 0.44 | 1.23 (0.70–2.14) | 0.47 | 1.06 (0.60–1.86) | 0.84 |
|  | **P for trend** |  |  | 0.55 |  | 0.58 |  | 0.92 |
| **Liver-related death** | Any CHIP | 412 | 1.26 (0.83–1.93) | 0.28 | 1.24 (0.81–1.89) | 0.32 | 1.20 (0.78–1.83) | 0.41 |
|  | Small CHIP |  | 1.34 (0.66–2.69) | 0.42 | 1.31 (0.65–2.64) | 0.45 | 1.49 (0.74–3.01) | 0.26 |
|  | Large CHIP |  | 1.23 (0.73–2.06) | 0.44 | 1.20 (0.72–2.02) | 0.48 | 1.08 (0.64–1.82) | 0.78 |
|  | **P for trend** |  |  | 0.37 |  | 0.41 |  | 0.56 |

**Model 1** adjusted for age, sex, and ethnicity; **Model 2** additionally adjusted for education level, Townsend deprivation index, smoking status, physical activity, daily alcohol intake, diet risk score, and BMI; **Model 3** (the fully adjusted model) further adjusted for prevalent diabetes, hypertension, statin use, and baseline levels of triglycerides, HDL cholesterol, CRP, ALT, GGT, and platelet count. P for trend was calculated using the ordinal CHIP category variable (0 = No CHIP, 1 = Small CHIP, 2 = Large CHIP).

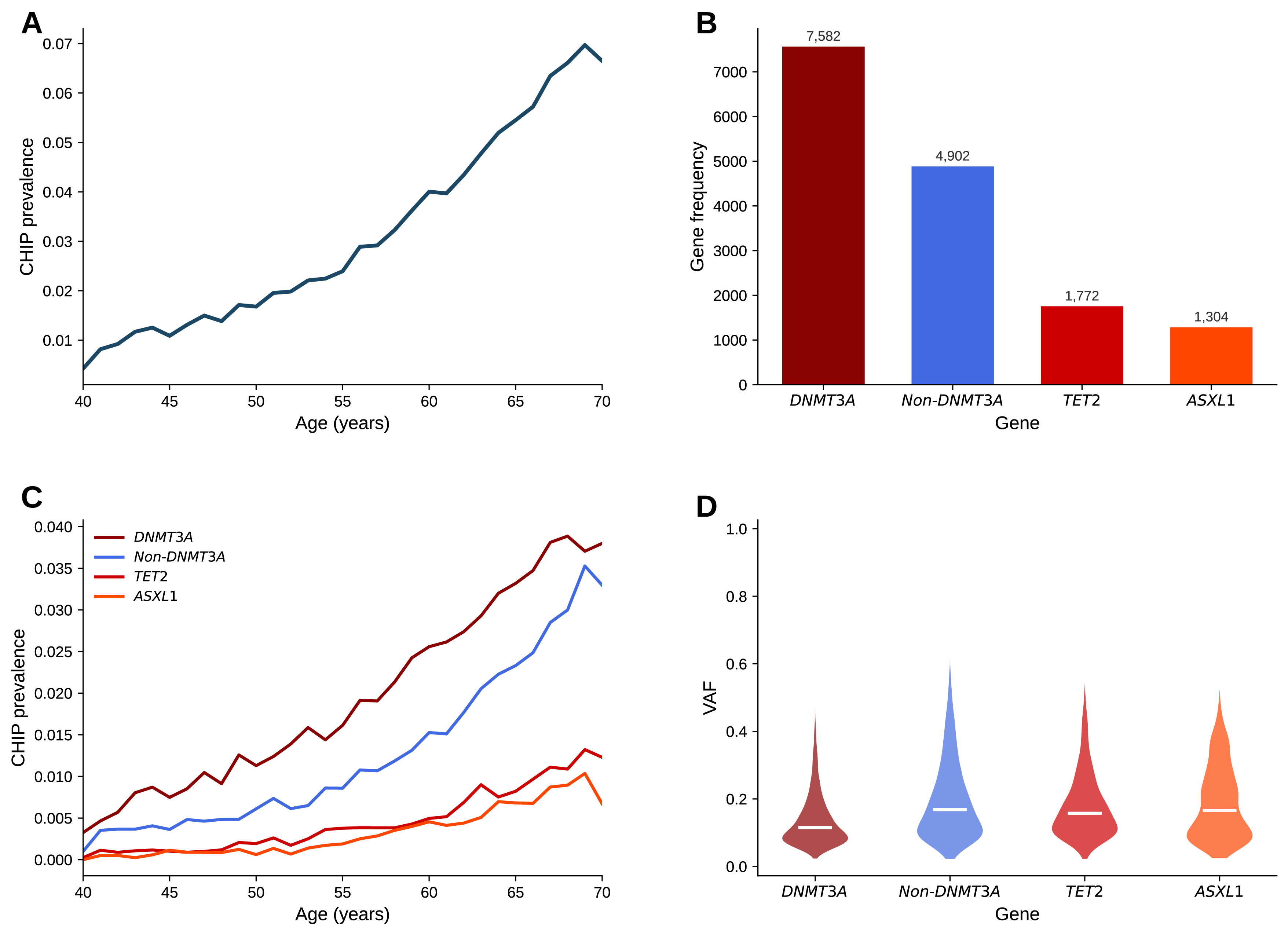

### **Supplementary Figure S1.** Prevalence of Clonal Hematopoiesis of Indeterminate Potential (CHIP) by Age Group

Bar plot displaying the prevalence of any CHIP, defined as a somatic mutation with a variant allele fraction (VAF) ≥2%, across age groups in the full study cohort (N=353,218). The prevalence of CHIP increases markedly with age, from less than 1% in individuals younger than 50 years to over 7% in those older than 70 years.

**Abbreviations:** CHIP, clonal hematopoiesis of indeterminate potential; VAF, variant allele fraction.

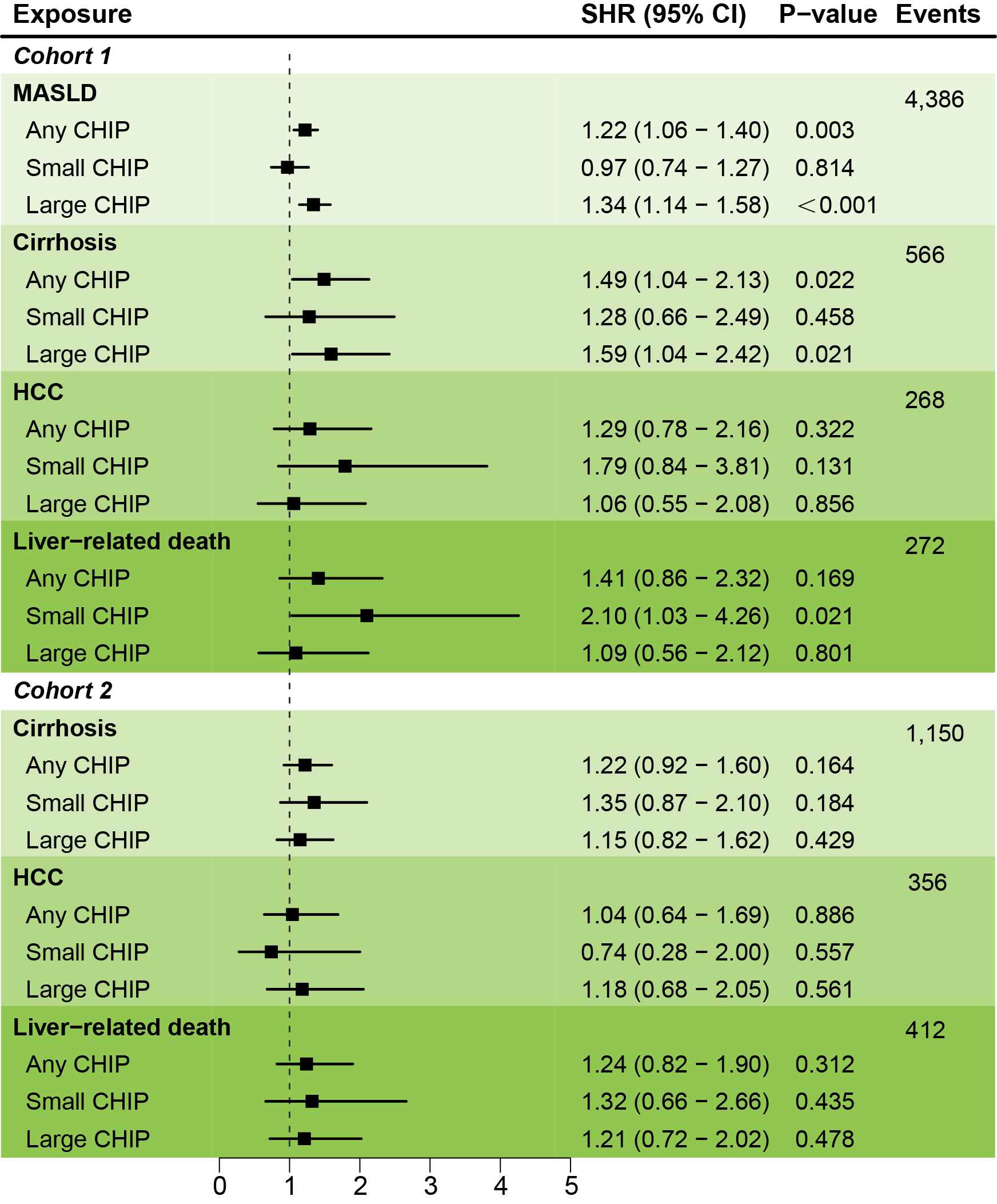

### **Supplementary Figure S2.** Competing Risk Analysis: Association Between CHIP and Liver Outcomes in the Healthy Population (Cohort 1)

Associations of any CHIP, CHIP stratified by clone size (Small: 2% ≤ VAF < 10%; Large: VAF ≥10%) with incident MASLD, cirrhosis, HCC, and liver-related death are shown. Models were fully adjusted for age, sex, ethnicity, education, Townsend deprivation index, smoking status, physical activity, daily alcohol intake, diet risk score, BMI, prevalent diabetes, hypertension, statin use, and baseline levels of triglycerides, HDL cholesterol, CRP, ALT, GGT, and platelet count.

**Abbreviations:** ALT, alanine aminotransferase; BMI, body mass index; CHIP, clonal hematopoiesis of indeterminate potential; CI, confidence interval; CRP, C-reactive protein; GGT, gamma-glutamyl transferase; HCC, hepatocellular carcinoma; HDL, high-density lipoprotein; MASLD, metabolic dysfunction-associated steatotic liver disease; SHR, subdistribution hazard ratio; VAF, variant allele fraction.

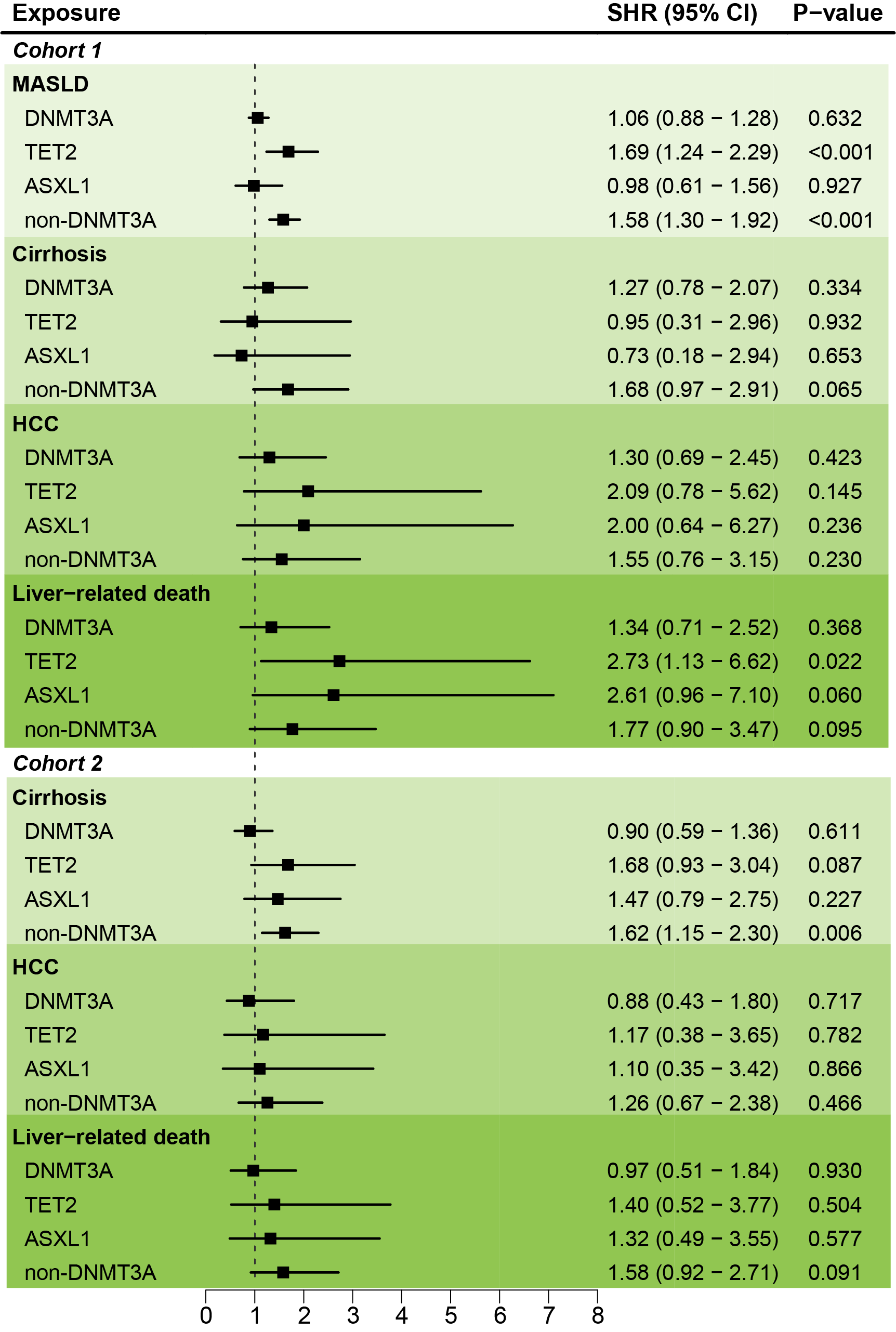

**Supplementary Figure S3.** Competing Risk Analysis: Association Between CHIP and Liver Disease Progression in the MASLD Population (Cohort 2)

Associations of CHIP stratified by driver gene (DNMT3A, TET2, ASXL1, non-DNMT3A) with incident cirrhosis, HCC, and liver-related death are shown. Models were fully adjusted as described in Supplementary Figure S2. **Abbreviations:** as in Supplementary Figure S2.

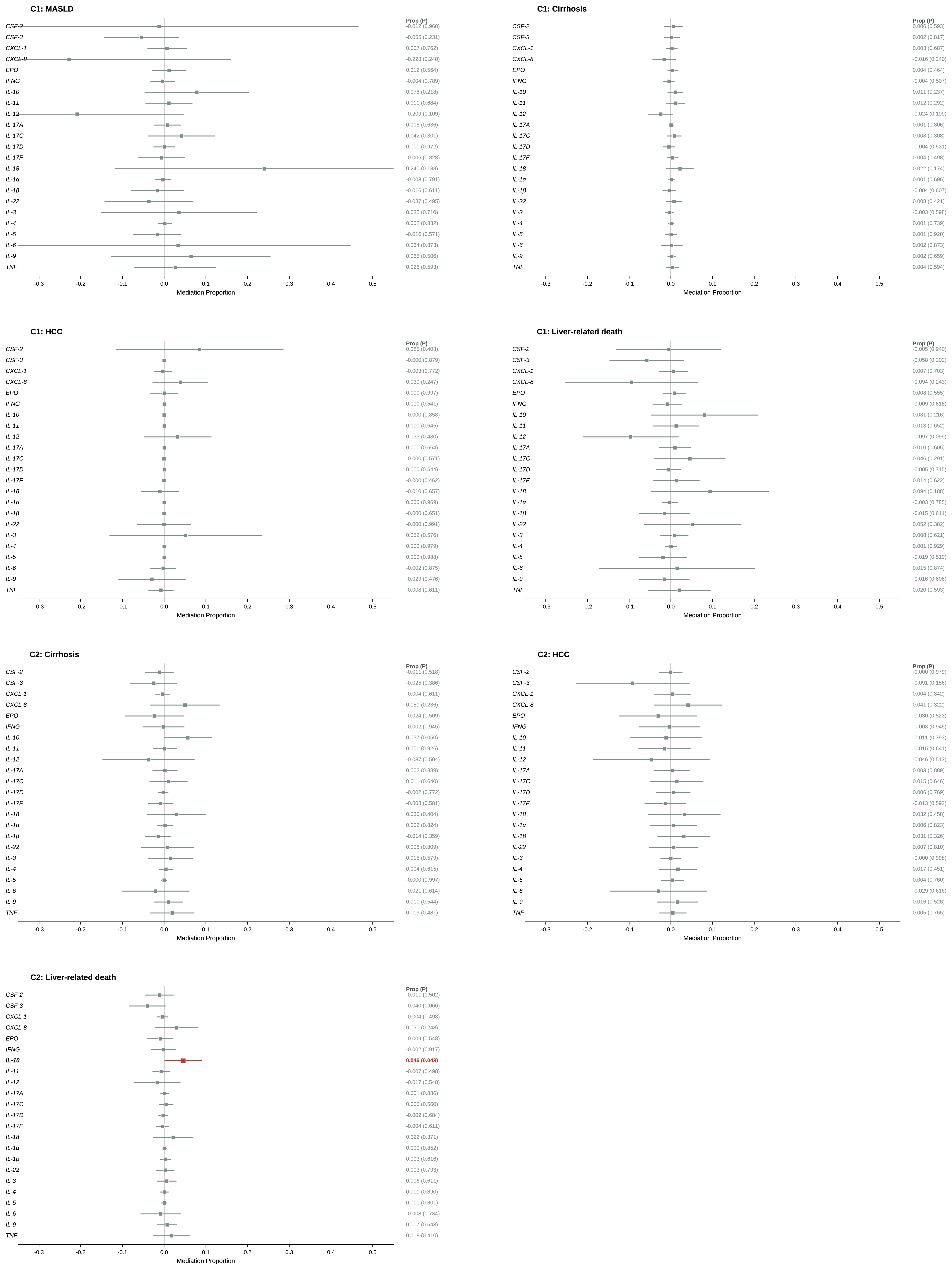

### **Supplementary Figure S4.** Mediation of the Association Between CHIP and Liver-Related Death by Circulating Cytokines (Olink Platform)

Forest plot showing the proportion of the total effect of CHIP on liver-related death in Cohort 2 that is mediated by each of 25 circulating cytokines measured using the Olink proximity extension assay. The proportion mediated and 95% confidence intervals were estimated using a causal mediation analysis framework with 1,000 bootstrap resamples. Models were adjusted for the full set of covariates as described in the Methods. Among the 25 cytokines tested, only interleukin-10 (IL-10) showed a statistically significant mediating effect.

**Abbreviations:** CHIP, clonal hematopoiesis of indeterminate potential; CI, confidence interval; IL, interleukin.
